# Long COVID risk by pre-infection symptoms and functional status: A retrospective cohort study of data from the *All of Us* Research Program

**DOI:** 10.1101/2025.08.07.25333259

**Authors:** Kristen Kehl-Floberg, Emma Freisberg, Aurora Pop-Vicas, Ron Gangnon, Dorothy Farrar Edwards

**Affiliations:** School of Medicine and Public Health, University of Wisconsin-Madison, Madison, Wisconsin, U.S.A; Department of Kinesiology, School of Education, University of Wisconsin-Madison, Madison, Wisconsin, U.S.A; Geriatric Research, Education, and Clinical Center (GRECC), William S. Middleton Veteran’s Memorial Hospital, Madison, Wisconsin, U.S.A; Berlin School of Public Health, Charité – Universitätsmedizin, Berlin, Germany

## Abstract

**IMPORTANCE:** Over seven million U.S. adults experience persistent health issues after COVID-19, known as “long COVID”. Although multiple guidelines recommend the inclusion of functional status in long COVID diagnostic criteria, more evidence is needed to guide this recommendation. This study explored the adjusted odds of developing long COVID by pre-infection symptoms and functional status, and the feasibility of estimating functional status using health records data.

**DESIGN & METHODS:** Retrospective cohort study of U.S. adults with history of COVID-19 enrolled in a multicenter national cohort study through July 2022 (*All of Us* Controlled Tier CDR 7.0), using diagnostic, procedure, and billing codes from the health record, and baseline survey responses. The risk of long COVID was estimated using logistic regression by pre-infection (ࢤ5 years) incidences of (a) at least one symptom common in long COVID, and (b) functional status, and adjusted for disease and demographic characteristics.

**RESULTS:** *N =* 65,464 met inclusion criteria; *n*=40,655 had post-infection occurrences of at least one symptom (long COVID group), *n=*24,809 had none (recovered). Adjusted odds ratios of developing long COVID increased with older age, female sex, Black racial identity, earlier variant, non-vaccination, lower pre-infection self-reported mental and cognitive health, and number of pre-infection symptoms. Adjusted odds were not significantly affected by any single pre-infection symptom, self-rated physical ability, or EHR-derived indicators of prior functional impairment.

**CONCLUSIONS:** In this model, there were no significant differences in risk of long COVID based on either pre-infection total incidences of long COVID symptoms (compared to the average of 4) or pre-infection functional impairment. This suggests that long COVID was associated with a change from baseline functioning and health, including in people with pre-infection incident symptoms and functional impairments. The impacts of co-occurring pre-infection symptoms requires further investigation. Both harmonized electronic health records data and patient-reported outcomes contribute important data for developing the diagnostic utility of functional status changes in long COVID.

## Introduction

Among U.S. adults with a history of SARS-CoV-2 infection, 7.7 to 23 million experience persistent post-acute symptoms [1] termed “long COVID” [2]. About 7.5% of U.S. adults reported long COVID symptoms in June 2024, and about 5.5% reported activity limitations as a result [3]. Long COVID is a patient-generated term for new-onset or exacerbated, and persistent, symptoms and health conditions after COVID-19 infection [2, 4–9]. Common presentations include fatigue, post-exertional malaise, cognitive impairment, headache, breathing difficulty, musculoskeletal pain or weakness, sleep disturbance, and mood disorder [10] among many others [11–13]. Long COVID symptoms affect complex and “chosen” activity patterns (such as productivity and leisure) more dramatically than basic self-care [14, 15] forcing some patients to abandon complex routines and roles in work, family, and community life [6, 16].

Clinical diagnostic definitions and criteria of long COVID are an urgent and ongoing focus of study [2]. Multiple research initiatives have been launched to study recovery from COVID-19 in large samples with the goal of identifying unique signatures of symptoms and biomarkers [11, 17–21], many using electronic health records (EHR) data. These studies have marshalled extraordinary amounts of data into various phenotypes and diagnostic hallmarks [22], providing vital direction on the disease and its impact. However, few have included quantifiable indicators of daily functioning [23], despite early patient reports describing profound impact on daily life [6, 24] and proposed criteria asserting that an impact on daily functioning is a critical diagnostic component [2, 25]. Because most EHR-based phenotyping studies have not included data on how disability manifests or is treated, their resulting diagnostic classifications have excluded this diagnostic feature.

Additionally, although post-infection presentations have been thoroughly investigated, studies of pre-infection occurrences of these same symptoms, conditions, and functional impairments are limited [26, 27]. Concerning symptoms, most studies that explore the emergence of symptoms and conditions after COVID-19 have not compared this to, nor assessed risk based on, their pre-infection prevalence [22]. Concerning functional status, pre-infection disability has been found to be associated with a two- [28] to three-fold [29] increase in long COVID, and pre-infection sense of well-being and mood have also been found to be associated with long COVID [30]. The meaning and utility of post-infection functional status as a diagnostic criterion requires evidence of the patterns of functional status that preceded infection, both in people who did and did not develop post-infection sequelae. Because long COVID includes worsened or exacerbated conditions that were previously well-managed [5], both pre-infection symptoms and functional status may be relevant to estimating risk of developing long COVID and tailoring management strategies [22].

The aim of this study is to explore whether pre-infection functional status and pre-infection occurrences of common long COVID symptoms are associated with development of at least one long COVID symptom, adjusted for demographics and infection characteristics, in a diverse population-based sample.

## Materials and Methods

### Design and data source

This is a retrospective cohort study of participants enrolled in the U.S. National Institutes of Health *All of Us* Research Program through July 2022 (curated data repository version 7.0), Controlled tier C2022Q4R9 [31] accessed through the *All of Us* Researcher Workbench [32] between May 1 2023 and March 3, 2026. *All of Us* is among the most comprehensive population-level biobanks in the world [33] enrolling participants from all 50 U.S. States and three territories and over-recruiting from groups that are historically underrepresented in medical research. Participants share baseline function and health data through surveys, biospecimens, and release of EHR data which is subsequently scraped every three months. The scientific question, approaches, and anticipated findings of the first author’s use of these data have been posted for review by participants and the public on the program’s Research Project Directory. The program and data snapshots can be viewed at https://www.researchallofus.org/data-tools/.

### Sample

*All of Us* participants 18 years of age and older with at least one incidence of any indicator of COVID-19 illness were included. Diagnostic indicators were (a) laboratory values for positive PCR, antigen, or antibody test, (b) self-reported illness assessed from May 2020 through February 2021 with the *All of Us* COVID-19 Participant Experiences (COPE) survey, or (c) the ICD-10 code for COVID-19 (U07.7) (S1 Appendix A, Table A.1). Informed consent for sharing EHR and survey data was obtained from all participants by *All of Us* Health Provider Organizations responsible for participant recruitment and enrollment. IRB oversight was delegated to the *All of Us* Central IRB. In compliance with *All of Us* data use agreement policies, investigators transformed date of birth to years of age promptly following import, and collapsed counts of fewer than 20 to prohibit subgroup membership identification in summary tables.

### *Long COVID* and *Recovered* participant groups

Participants with COVID-19 history were grouped by whether their EHR showed at least one diagnosis or observation code from a list of 38 long COVID symptom and condition categories (see below, “Pre- and post-infection incidences of long COVID symptoms”, and S1 Appendix A, Table A.2) at least 28 days after first infection. Participants with at least one symptom were classified in the “long COVID” group; the rest were classified in the “recovered” group. The post-infection interval of 28-days was chosen to capture health and daily functioning beginning after the presumptive acute infection phase but still within the 90-day timeframe of medical leave allowable under the U.S. law [34]. While this is sooner post-infection than the 90-day window in other studies [22] and the WHO proposed diagnostic criteria [25], a shorter interval is of interest to both classification and clinical interests for targeted rehabilitation services. Because functional impairment is proposed as part of the diagnostic classification, we wished to characterize long COVID in a timeframe that would be clinically useful in the U.S. for utilizing skilled rehabilitation for functional impairment. (S1 Appendix A).

### Data sources and variables

#### Surveys: Demographics, self-reported health, and self-reported function at study enrollment

*All of Us* participants provide self-report of their health and daily functional status data through validated self-report measures from the PROMIS battery and *All of Us*-designed surveys at enrollment. This study used responses to “The Basics” (demographics), “Overall Health” (daily functioning and mental/cognitive health) survey from the PROMIS Two-Item Global Health surveys [35] and the “Covid-19 Participant Experience” (COPE) (self-reported COVID-19 symptoms between May 2020 – Feburary 2021) surveys.

#### Pre- and post-infection incidences of long COVID symptoms and conditions

Symptoms of long COVID were chosen a-priori based on the Centers for Disease Control and Prevention’s list of symptoms [9] and emergency billing code for Post-COVID conditions [36], prior phenotyping and patient-led literature, and the authors’ clinical and community research experience. These sources yielded a list of 44 symptoms/conditions (“descendent” codes), which fell under 38 categories (“ancestor” codes) within the *All of Us* data table structure (S1 Appendix A, Table A.2). These 38 ancestor diagnoses were queried for all participants. For each category, the first incidence in the five years prior to infection signified a pre-infection occurrence, and the first incidence 28 days or more after the infection signified a post-infection occurrence (long COVID group only) of any symptom under that category. Because proposed empirical definitions of long COVID include exacerbations or relapses of pre-infection conditions [5, 13, 22], we queried “any mention” of the symptoms/conditions in this list (rather than “first mention”). Symptom persistence attributable to COVID-19 infection was not estimated due to the *All of Us* data structure and privacy policies. S1 Appendix A reports selection process, rationale, condition categories, and exclusions.

#### Total number of long COVID symptoms/conditions present prior to infection

From the pre-infection symptom occurrences, a gross measure of pre-infection symptom burden was computed for each participant by tallying all long COVID symptom categories with observations. This included all symptoms (e.g. headache, joint pain, cough) and conditions (ME/CFS, POTS, and post-viral fatigue syndromes) used in cohort discovery.

#### Infection variants active at time of first infection and vaccination status

A time-bound variable was created to infer variant exposure at date of first SARS-CoV-2 infection. Periods began and ended around the emergence of major variants as reported by Markov et al [37]. Our variant periods were “pre-VOC/wild type” from January-November 2020, “pre-VOC/alpha/beta from November 2020 – April 2021, “alpha/beta/delta” from April-August 2021, “delta” from August-December 2021, “omicron_BA1-BA2” from December 2021 – April 2022, and “omicron_BA2-BA5” from April 2022 – July 2022 (data release cut-off date).

Vaccination was noted as “full series” or “not vaccinated” according to the CDC’s definition at the time of data cut-off (at least a two-dose primary series of mRNA or at least one dose of all other types) [38] (Appendix A, Table A.3).

#### Demographics

Age, sex assigned at birth, race, and ethnicity were included based on evidence that these factors influence risks for both long COVID [39–42] and functional impairment [43, 44]. Level of education was included as a proxy for social determinants of health due to its impact on access to health-supporting societal resources [45], as financial concerns [46] and greater unmet social needs [47] have been found to be associated with more numerous and persistent long COVID symptoms.

#### Pre-COVID daily functioning

Pre-infection functional status was explored using enrollment surveys and functional status findings (EHR procedure and diagnostic codes) recorded between January 1, 2015 and four weeks prior to first infection. Survey responses were queried for self-reported performance of physically demanding daily activities, social roles, and mental/cognitive health. From the EHR, the incidences of medical billing codes for therapeutic procedures provided by occupational therapists were tallied [48] (S1 Appendix B). Additionally, procedure codes under the vocabulary hierarchy “Finding of Functional Performance and Activity” were grouped into three levels of presumable impairment (“None”, “Some”, and “Dependent on others”). (S1 Appendix B.)

### Quantitative bias analysis of cohort discovery

Potential cohort bias was examined using non-parametric propensity score matching.

Bias was estimated for two possible classification-based approaches: (a) participant enrollment before first COVID infection (retained for this analysis) versus after (excluded), and (b) a 90 day symptom start date instead of 28 days (S1 Appendix C).

### Analysis

Demographics of the long COVID and recovered groups were graphed and tested for significant difference using Welch’s *t* test (normal), Mann-Whitney U test (non-normal), or *X*^2^ test (categorical). Unadjusted odds ratios were computed for all variables. Logistic regression was used to model the adjusted odds of developing long COVID based on pre-infection symptoms/conditions and pre-infection functional status, adjusted for demographics, variant, and vaccination status (S1 Appendix D, Table D.1-D.2, Fig D.1). Data access and analyses were performed in the *All of Us* Researcher Workbench, Jupyter notebooks cloud computing environment, using R [49]. Cohort discovery, data cleaning and organization, and statistical analyses were completed by the first author using R packages “stats” [50], “rms” [51], “corrplot” [52], and the “tidyverse” suite [53].

## Results

104,993 participants met inclusion criteria of at least one COVID-19 illness. Participants with missing demographic survey responses (*n*=21,202) were removed. Further removed were *n*=16,815 participants who enrolled after their first COVID-19 indicator (allowing use of enrollment questionnaires as pre-infection self-reported baseline functioning and health data); *n*=1,506 due to non-responses to the social survey questions (permitting these responses to be combined as an ordinal summary scale); *n=*5 participants with erroneous diagnostic codes (concept name and code was for SARS-CoV-2, but the dates of diagnosis corresponded with previous SARS-type virus outbreaks in the early 2000’s); and *n*=1 due to an extreme and clinically unlikely value of 1286 (or 321 hours) for their 5-year CPT billing code total. The final sample was *N =* 65,464 participants, with 40,655 showing at least one post-infection symptom (long COVID group) and 24,809 showing none (recovered group) (Fig 1).

**Fig 1.**
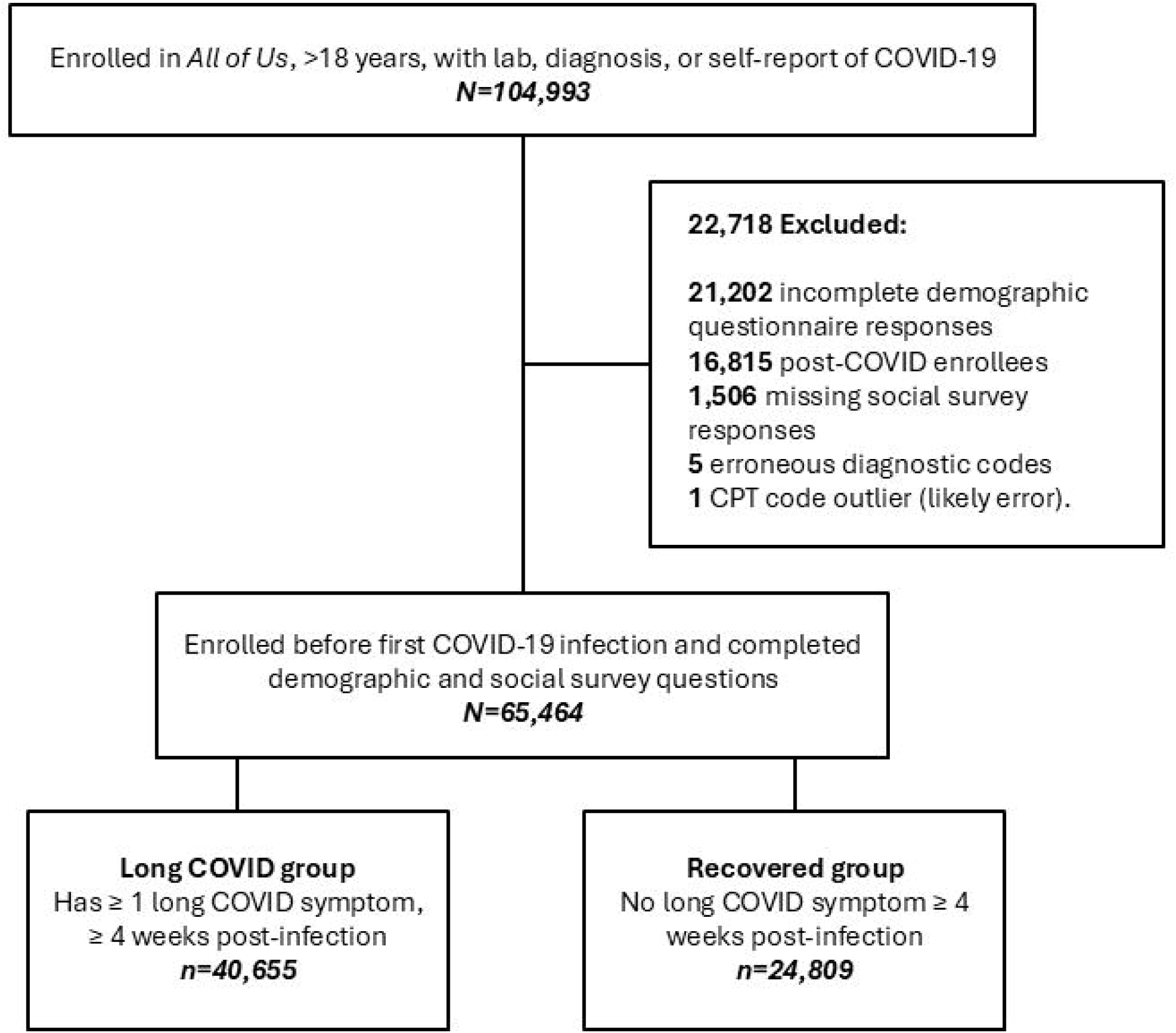
Diagram of the inclusion of participants for final analysis from the available sample of *All of Us* participants, curated data repository v7, data through July 1, 2022.

### Quantitative bias analysis of cohort discovery

When comparing pre- and post-infection enrollees, non-parametric propensity score matching showed a low match rate of 45% between pre- and post-infection enrollees (S1 Appendix C, Figs C.1-C.2) suggesting that more recent enrollees (excluded from this analysis) differed in some demographic and disease characteristics. This may have been related to programmatic influences on recruitment that affect the sample over time such as adjustments in recruitment to increase sample diversity or the addition of new study sites in previously under-represented regions or patient populations, and may indicate that some of these groups are underrepresented in this sample. When classifying participants into long COVID groups using a 90-day delay from first infection, 7.2% (n=3,032) participants who had been cases at the-28 day classification period moved into the control group, leaving n=38,607 long COVID cases. As none of the original controls were re-classified under the longer lag time, this group showed no differences. The standardized mean differences between cases and controls using this study’s original 28 day classification timeframe remained substantial (above 0.1). This indicates that use of a 90-day classification scheme would have changed the group distributions very little. A 28-day classification of long COVID symptoms was thus retained for these analyses. (S1 Appendix C, Figs (C.3-C.4).

### Model fit

The final model’s moderate *R^2^* (0.449) accounted for just under 50% of the total variance. This was not suspected to be due to bias, as the regularity of the residual plots were satisfactory (S1 Appendix D). Multicollinearity among all variable parameters in the model was assessed using variance inflation factor (VIF) (Appendix D). No parameters exceeded the chosen threshold of 10. VIF values over 5 were observed for some control variables (Age (6.1), and Black (5.89) or White race (6.16)). For the primary predictors, variance was slightly inflated for pre-infection functional status (3.5-3.9), and more so for seven of the pre-infection symptoms’ incidences (5.3-9.5; see Appendix D, Table D.4). Symptoms’ high VIF may have been caused by their inclusion in the total number of pre-infection incident symptoms, which had a VIF score of 6.9 for at least one symptom. The interaction between number of pre-infection symptoms and years of age was also high (7.29-8.84 for lower numbers of symptoms). These multiplicative and redundant variables were expected to be collinear with their component variables.

### Demographics, variant, and vaccination status

Demographic distributions, and their unadjusted odds ratios and multivariate adjusted odds ratios, are shown in Table 1. Most participants had their first infection during the pre-variant/wild type period, during which the proportion of long COVID participants outnumbered that of recovered participants by almost 2 to 1 (S1 Appendix E, Fig E.1). The Long COVID participants were older (median 63 (24) versus 60 (28)), and had slightly higher percentages of female and Black participants than the recovered group. Groups were effectively equivalent in numbers of participants reporting Hispanic/Latino ethnicity. Adjusted odds of developing long COVID after infection increased with older age. Participants with female biological sex and Black or African American racial identity showed slightly higher odds. Adjusted odds were also higher with earlier variant at first infection, decreasing over each subsequent variant period. Non-vaccination with full primary series was associated with 48% (1.29, 1.69) higher odds (*p*<0.000). The small 4.9% proportion of this sample that had received any combination of vaccines considered to be the full vaccination series by data cut-off resulted in sparse and unevenly distributed counts when stratified by the primary factors. The relative timing of vaccination and variant exposures are graphed in S.1. Appendix E, Fig E.2.)

**Table 1.** Demographic, functioning, and long COVID disease characteristics and associated odds ratios of developing long COVID in *All of Us* participants through July 2022.

|  | Long COVID Symptoms | Recovered | *P* | OR | AOR (99% CI), *P* |
| --- | --- | --- | --- | --- | --- |
| *N* = 65464 | *n*=40655 | *n*=24809 |  |  |  |
| Age (mean (SD)) | 61.07 (16.03) | 57.73 (16.81) | < 2.2e-16 |  |  |
| Age (median (IQR)) | 63 (24) | 60 (28) | < 2.2e-16 |  |  |
| 25 |  |  |  | 0.54 (0.43, 0.69), <0.000 | 0.68 (0.60, 0.78), <0.000 |
| 35 |  |  |  | 0.64 (0.59, 0.70), <0.000 | 0.76 (0.69, 0.83), <0.000 |
| 45 |  |  |  | 0.73 (0.68, 0.79), <0.000 | 0.84 (0.79, 0.89), <0.000 |
| 55 |  |  |  | 0.94 (0.87, 1.02) 0.06 | 0.93 (0.91, 0.95), <0.000 |
| 62 |  |  |  | 1 | 1 |
| 75 |  |  |  | 1.09 (1.02, 1.17), <0.000 | 1.14 (1.09, 1.20), <0.000 |
| 85 |  |  |  | 1.21 (1.12, 1.31), <0.000 | 1.27 (1.17, 1.38), <0.000 |
| Sex assigned at Birth (%) |  |  | < 2.2e-16 |  |  |
| Female | 27230 (65.4) | 15571 (61.5) |  | 1.18 (1.13, 1.2), <0.000 | 1.08 (1.02, 1.14), <0.000 |
| Male | 14403 (34.6) | 9754 (38.5) |  | 1 | 1 |
| Race (%) |  |  | < 2.2e-16 |  |  |
| Asian | 989 (2.4) | 999 (3.9) |  | 0.62 (0.55, 0.69), 0.000 | 0.92 (0.79, 1.1), 0.14 |
| Black or African American | 9311 (22.4) | 4822 (19.0) |  | 1.20 (1.14, 1.26), 0.000 | 1.12 (1.04, 1.2), <0.0 |
| Middle Eastern or North African | 293 (0.7) | 201 (0.8) |  | 0.89 (0.70, 1.13), 0.21 | 1.03 (0.76, 1.4), 0.78 |
| More than one population | 805 (1.9) | 536 (2.1) |  | 0.93 (0.80, 1.07), 0.93 | 1.01 (0.84, 1.2), 0.91 |
| White | 30241 (72.6) | 18774 (74.1) |  | 1 | 1 |
| **Ethnicity (%)** |  |  | 0.0002 |  |  |
| Hispanic or Latino | 458 (2.3) | 701 (2.8) |  | 0.83 (0.73, 0.94),<br><0.000 | 0.88 (0.74, 1.03), 0.035 |
| **Education (%)** |  |  | <2.2e-16 |  |  |
| Advanced Degree | 9722 (23) | 7230 (29) |  | 0.79 (0.76, 0.83),<br><0.000 | 0.97 (0.91, 1.03), 0.197 |
| College or college grad | 21425 (52) | 12661 (51) |  | 1 | 1 |
| High School/GED | 7043 (17) | 3658 (14) |  | 1.14 (1.07, 1.21),<br><0.000 | 0.96 (0.89, 1.03), 0.131 |
| Some High School | 1592 (4) | 851 (3) |  | 1.11 (0.98, 1.24), 0.02 | 0.85 (0.73, 0.98), 0.003 |
| Less than high school | 328 (<1) | 143 (<1) |  | 1.35 (1.05, 1.76),<br>0.002 | 0.97 (0.70, 1.34), 0.798 |
| No Answer | 545 (1) | 266 (1) |  | 1.21 (0.99, 1.47), 0.01 | 1.03 (0.81, 1.32), 0.725 |
| **Variant of Concern** |  |  | < 2.2e-16 |  |  |
| Pre-VOC/wild type | 21632 (52.0) | 7575 (29.9) |  | 1 | 1 |
| Pre-VOC, alpha, and beta | 10138 (24.3) | 6446 (25.4) |  | 0.55 (0.52, 0.58),<br><0.000 | 0.61 (0.57, 0.65), <0.000 |
| alpha, beta, and delta | 3317 (8.0) | 1922 (7.6) |  | 0.60 (0.55, 0.65),<br><0.000 | 0.48 (0.44, 0.53), <0.000 |
| delta | 3538 (8.5) | 2945 (11.6) |  | 0.42 (0.39, 0.46),<br><0.000 | 0.32 (0.29, 0.35), <0.000 |
| omicron BA1-BA2 | 2629 (6.3) | 3615 (14.3) |  | 0.25 (0.24, 0.27),<br><0.000 | 0.17 (0.15, 0.18), <0.000 |
| omicron BA2-BA5 | 385 (0.9) | 2829 (11.2) |  | 0.05 (0.04, 0.05),<br><0.000 | 0.02 (0.02, 0.02), <0.000 |
| **Vaccination with full primary series(1) (%)** |  |  | < 0.001 |  |  |
| Vaccinated | 2415 (5.9) | 795 (3.2) |  | 1 | 1 |
| Not Vaccinated | 38240 (94.1) | 24014 (96.8) |  | 1.91 (1.71, 2.13),<br><0.000 | 1.48 (1.29, 1.69), <0.000 |

Table 1 Legend: Distributions, central tendency, spread, unadjusted odds ratios and adjusted odds ratios of sample demographics, variant at first infection, and vaccination status.

### Pre-infection daily functioning

EHR-recorded billing code indicators of daily functioning showed higher proportions of pre-infection performance difficulty (10% vs. 4%) and dependence on others for care (6% vs. 2%) in the long COVID group. In baseline functional surveys, participants in the long COVID group more frequently selected lower ratings on physical activity ability, social score, and mental health. However, when adjusted for demographics and pre-infection symptom occurrences, a “good”, or moderate, level of self-rated mental/cognitive health was the only indicator that was associated with increased odds of developing long COVID (Table 2). This was also evident in the interactions between these indicators and age; neither self-rated nor EHR-derived physical independence was a significant predictor of long COVID at any age (S1 Appendix E, Tables E.1 – E.2). (An interaction between age and self-reported mental health was not run due to indications of poor model fit with this term (S1 Appendix D).)

**Table 2.** Pre-infection daily functioning and associated odds ratios of developing long COVID in *All of Us* participants through July 2022.

|  | Long<br>COVID<br>Symptoms | Recovered | OR | AOR (99% CI), *P* |
| --- | --- | --- | --- | --- |
| *N*=65464 | *n*=40655 | *n*=24809 |  |  |
| **Pre-infection functional**<br>**performance difficulty (%)** |  |  |  |  |
| None | 33933<br>(83.5) | 23494<br>(94.7) | 1 | 1 |
| Some | 4106 (10.1) | 926 (3.7) | 3.07 (2.79, 3.38), <0.000 | 1.01 (0.63, 1.63), <0.9585 |
| Severe or dependent | 2616 (6.4) | 389 (1.6) | 4.65 (4.05, 5.38), <0.000 | 1.41 (0.85, 2.31), <0.078 |
| **Total CPT units billed 5yrs-28 days prior to**<br>**infection** |  |  |  |  |
| mean(sd) | 4.45 (17.78) | 2.06<br>(10.22) |  |  |
| 0 | 30325<br>(77.1) | 21282<br>(85.8) | 1 | 1 |
| 5 | 4263 (10.5) | 1618 (6.5) | 1.85 (1.71, 2.00), <0.000 | 1.00 (0.99, 1.0), 0.23 |
| 20 | 3601 (8.9) | 1183 (4.8) | 2.14 (1.95, 2.34), <0.000 | 0.98 (0.94, 1.0), 0.23 |
| 80+ | 2138 (5.3) | 656 (2.6) | 2.29 (2.04, 2.57), <0.000 | 0.93 (0.79, 1.1), 0.23 |
| **To what extent are you able to carry out your everyday physical activities?** |  |  |  |  |
| Completely | 21470 (52.8) | 16608 (66.9) | 1.00 | 1.000 |
| Mostly | 7715 (19.0) | 3821 (15.4) | 1.56 (1.47, 1.65), <0.000 | 1.04 (0.97, 1.13), <0.140 |
| Moderately | 6415 (15.8) | 2542 (10.2) | 1.95 (1.83, 2.09), <0.000 | 1.02 (0.93, 1.12), <0.590 |
| A little | 4123 (10.1) | 1451 (5.8) | 2.20 (2.02, 2.39), <0.000 | 0.94 (0.84, 1.06), <0.211 |
| Not at all | 661 (1.6) | 254 (1.0) | 2.01 (1.66, 2.44), <0.000 | 0.83 (0.65, 1.06), <0.050 |
| No Answer | 271 (0.7) | 133 (0.5) | 1.58 (1.20, 2.08), <0.000 | 1.00 (0.71, 1.41), <0.998 |
| **In general, how would you rate your mental health, including your mood and your ability to think?** |  |  |  |  |
| Excellent | 8057 (19.8) | 6092 (24.6) | 1.00 | 1.000 |
| Very Good | 13078 (32.2) | 8950 (36.1) | 1.10 (1.04, 1.17), <0.000 | 1.04 (0.96, 1.12), 0.235 |
| Good | 11924 (29.3) | 6286 (25.3) | 1.43 (1.35, 1.52), <0.000 | 1.14 (1.04, 1.25), <0.000 |
| Fair | 6009 (14.8) | 2781 (11.2) | 1.63 (1.52, 1.76), <0.000 | 1.02 (0.91, 1.15), 0.612 |
| Poor | 1279 (3.1) | 545 (2.2) | 1.77 (1.55, 2.04), <0.000 | 1.11 (0.90, 1.36), 0.206 |
| No Answer | 308 (0.8) | 155 (0.6) | 1.50 (1.16, 1.95), <0.000 | 0.97 (0.70, 1.33), 0.776 |

### Pre-infection occurrences of common long COVID symptoms and conditions

The range of pre-infection occurrences of common long COVID symptoms was between 0 and 23. The long COVID group had a higher median number of pre-infection symptoms (6(6) vs. 1(4), Table 3). The proportions of the number of participants with any observations under these broad categories prior to infection are shown in Fig 2 and Table 3. There were fewer than 500 incidences of anosmia with sparse cells by group, so this symptom was not included in the model. There were no pre-infection observations of POTS, PVFS, or ageusia.

**Table 3.** Pre-infection occurrence of common long COVID symptoms, and association with development of long COVID.

|  | **Long**<br>**COVID**<br>**Symptoms** | **Recovered** | **OR** | **AOR (99% CI), *P*** |
| --- | --- | --- | --- | --- |
| Total number of pre-pandemic symptoms of long COVID (mean (SD)) | 6.11 (3.96) | 2.45 (2.94) |  |  |
| Total number of pre-pandemic symptoms of long COVID (median (IQR)) | 6 (6) | 1 (4) |  |  |
| 0 |  |  | 0.13 (0.12, 0.14), <0.000 | 0.15 (0.04, 0.62), 0.009 |
| 1 |  |  | 0.47 (0.44, 0.51), <0.000 | 0.39 (0.13, 1.13), 0.083 |
| 4 |  |  | 1.00 | 1.00 |
| 7 |  |  | 2.13 (1.99, 2.27), <0.000 | 1.58 (0.55, 4.59), 0.398 |
| **Long COVID symptom/condition group** |  |  |  |  |
| Joint Pain | 25675 (62) | 7626 (30) | 3.74 (3.58, 3.91), 0.00 | 1.32 (0.83, 2.12), 0.12 |
| Chest Pain | 19400 (47) | 5054 (20) | 3.20 (3.34, 3.68), 0.00 | 1.03 (0.65, 1.65), 0.86 |
| Abdominal | 18803 (45) | 5237 (21) | 3.17 (3.02, 3.33), 0.00 | 1.07 (0.67, 1.71), 0.72 |
| Sleep | 18241 (44) | 3902 (15) | 4.29 (4.07, 4.52), 0.00 | 1.53 (0.96, 2.46), 0.02 |
| Anxiety | 17717 (43) | 4148 (16) | 3.80 (3.61, 3.99), 0.00 | 1.36 (0.85, 2.19), 0.09 |
| Depression | 16639 (40) | 3634 (14) | 3.99 (3.79, 4.21), 0.00 | 1.31 (0.82, 2.10), 0.14 |
| Cough | 15833 (38) | 3977 (16) | 3.31 (3.15, 3.49), 0.00 | 1.09 (0.68, 1.74), 0.64 |
| Dyspnea | 14670 (35) | 3308 (13) | 3.64 (3.44, 3.45), 0.00 | 1.14 (0.71, 1.82), 0.48 |
| Fatigue | 14172 (34) | 3339 (13) | 3.42 (3.23, 3.61), 0.00 | 1.13 (0.70, 1.81), 0.51 |
| Rash/Skin/Hair | 11488 (28) | 2965 (12) | 2.86 (2.70, 3.04), 0.00 | 1.27 (0.79, 2.04), 0.19 |
| Headache | 11051 (27) | 2603 (10) | 3.16 (2.97, 3.36), 0.00 | 0.99 (0.62, 1.59), 0.97 |
| Dizziness | 10388 (25) | 2398 (9) | 3.19 (3.00, 3.40), 0.00 | 1.04 (0.65, 1.67), 0.83 |
| Diarrhea | 9273 (22) | 2076 (8) | 3.22 (3.01, 3.45), 0.00 | 1.07 (0.67, 1.72), 0.70 |
| Muscle Pain | 6528 (16) | 1253 (5) | 3.60 (3.31, 3.92), 0.00 | 1.16 (0.72, 1.87), 0.41 |
| Palpitations | 6308 (15) | 1469 (6) | 2.91 (2.69, 3.15), 0.00 | 1.15 (0.71, 1.85), 0.45 |
| Fever | 5537 (13) | 1390 (5) | 2.63 (2.45, 2.88), 0.00 | 0.90 (0.56, 1.44), 0.55 |
| Cognition | 5131 (12) | 1117 (4) | 3.03 (2.78, 3.32), 0.00 | 1.01 (0.63, 1.64), 0.94 |
| Paraesthesia | 5111 (12) | 1169 (5) | 2.88 (2.64, 3.14), 0.00 | 1.09 (0.68, 1.76), 0.64 |
| Tachycardia | 4946 (12) | 1225 (5) | 2.68 (2.46, 2.92), 0.00 | 0.86 (0.53, 1.38), 0.40 |
| Menstrual Disorder | 4304 (10) | 1475 (6) | 1.87 (1.73, 2.03), 0.00 | 1.07 (0.67, 1.73), 0.70 |
| Musculoskeletal Chest Pain | 3220 (8) | 648 (3) | 3.20 (2.87, 3.60), 0.00 | 1.09 (0.67, 1.78), 0.64 |
| ME/CFS (Chronic Fatigue) | 2228 (5) | 430 (2) | 3.33 (2.90, 3.83), 0.00 | 1.04 (0.63, 1.72), 0.83 |
| Sexual Dysfunction | 847 (3) | 231 (1) | 2.26 (1.86, 2.75), 0.00 | 0.99 (0.60, 1.68), 0.99 |

**Fig 2.**
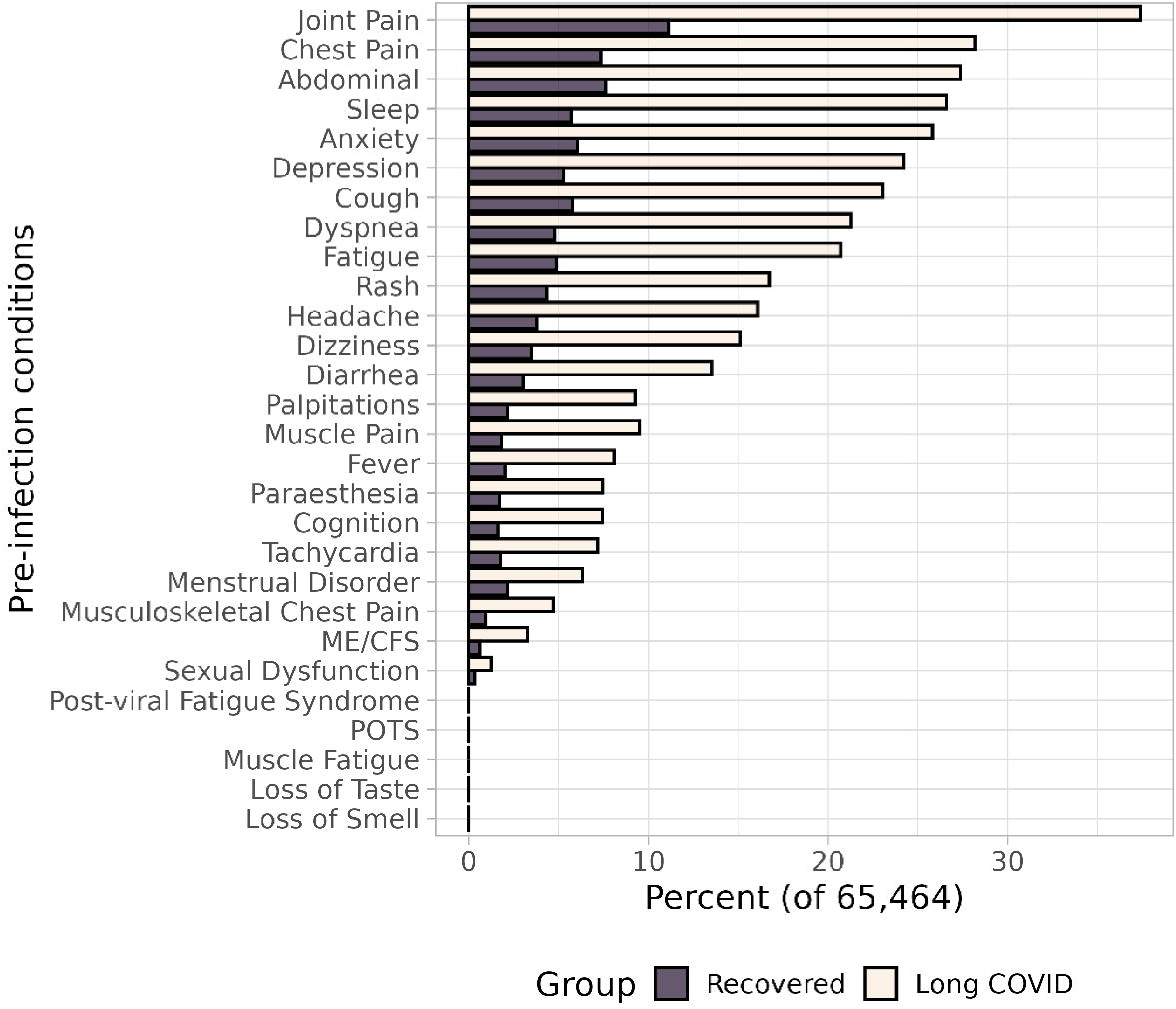
Proportion of each group with pre-infection incidences of common long COVID symptoms. A histogram with side-by-side light (long COVID group) and dark (recovered group) bars. Shows the differences in the proportion of each group experiencing each long COVID symptoms *before* infection (between 5 years and 28 days). The *Y* axis plots the symptoms or conditions from most-frequently observed (joint pain) to least (sexual dysfunction); the *X* axis plots the proportion with any observation of a symptom.

Table legend: Percent of participants with versus without long COVID with at least one pre-infection occurrence of common long COVID symptoms, with unadjusted and adjusted odds ratios of developing long COVID given each symptom/condition.

Compared to the median number of four pre-infection occurrences of any common long COVID symptom/condition, having zero incidences was associated with 85% lower adjusted odds of long COVID (*p*=0.009), with no significant differences for higher symptoms totals. This was also observed across ages in interactions between symptom totals and age (S1 Appendix E, Table E.3). Long COVID cases showed between three to five times greater prevalence of every pre-infection symptom/condition (Fig 2).None of these symptoms were significant predictors of long COVID in the adjusted model (Table 3) (sleep disturbance was the only symptom category that approached significance (*p*=0.02)), however ME/CFS, joint pain, and several others demonstrated high variance inflation factors (S1 Table D.4) indicating possible underestimation of their effects (S1 Appendix D).

Pearson’s *R* correlations were then computed to assess the pre-infection symptom categories for correlation (S1 Appendix E, Table E.4) and grouped by first principal components analysis (Fig 3). No strong clusters of symptoms were observed, supporting fitting symptoms individually. Depression and anxiety had the highest correlation (*r*=0.53). Moderate to low correlations were found between abdominal pain and both diarrhea (*r*=0.32) and chest pain (*r*= 0.32), dyspnea with both chest pain (*r*= 0.4) and cough (*r*=0.33), ME/CFS and fatigue (*r*=0.34), musculoskeletal and generic chest pain (*r*=0.33), and sleep and depression (0.31).

**Fig 3.**
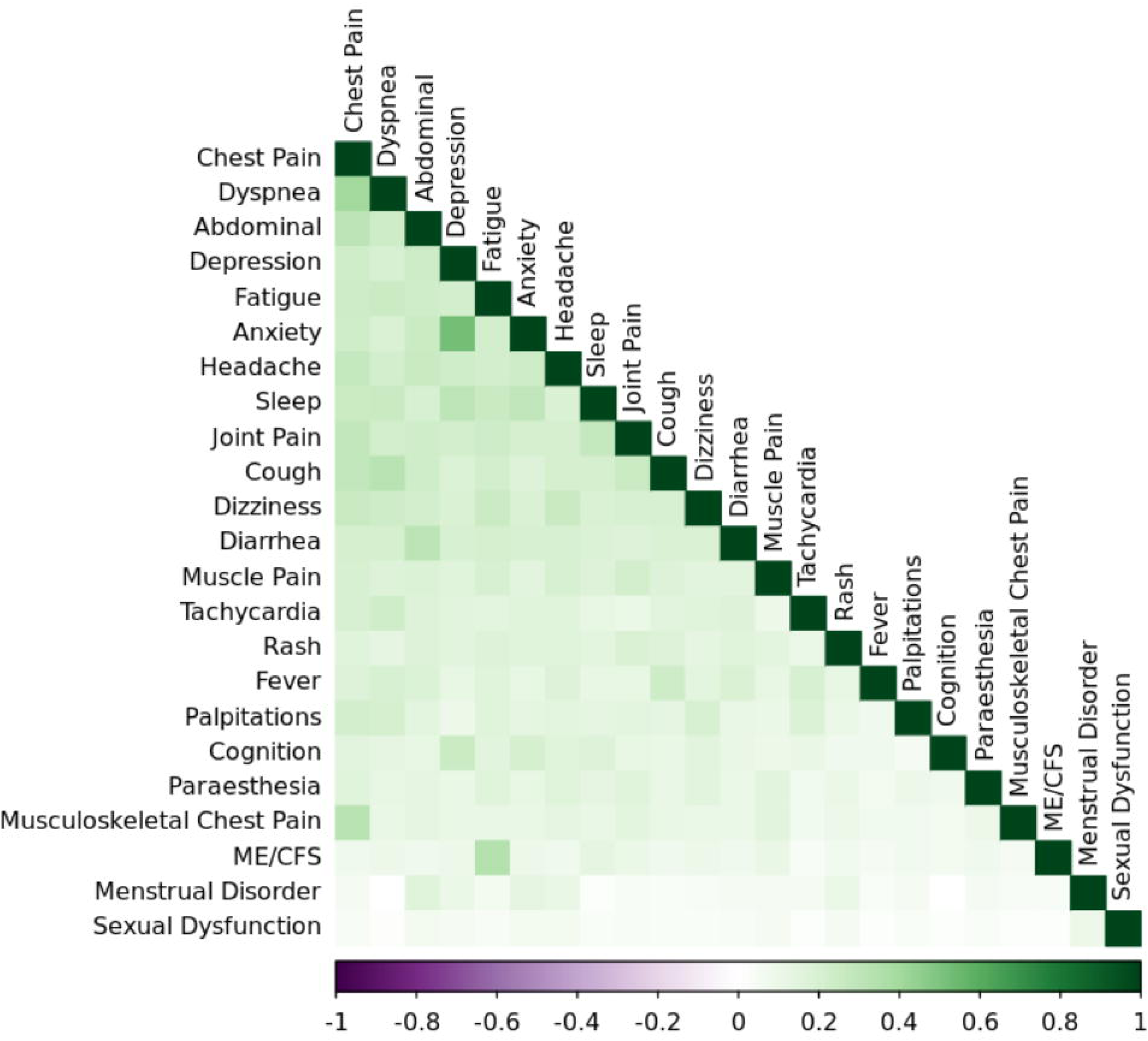
Pre-infection symptom/condition correlations grouped by first principal components analysis. Heat map of first principal components analysis of pre-infection symptom correlations reported in Appendix E Table E.4. Green colors indicate positive correlations (maximum = moderate correlation between anxiety and depression, *R*=0.53) and purple indicate negative correlations. Darker colors indicate stronger correlation, whereas paler colors indicate weaker.

White indicates no correlation (menstrual disorder with dyspnea, *R*=0.00). This heatmap shows a lack of clear and distinct groupings of pre-infection symptoms.

## Discussion

This retrospective cohort study is among the first examinations of long COVID among participants in the *All of Us* Research Program. To our knowledge, it is also the first population-level study to explore the prevalence of pre-infection occurrences of long COVID symptoms and functional status levels, and their associations with developing post-COVID symptoms.

These associative findings of pre-infection status contribute several insights into the evolving understanding of the risks of long COVID. Having a greater number of incident long COVID symptoms before infection, and having poorer functional status, were both associated with significantly higher unadjusted odds. However, in the full model with all covariates held at their most-frequently observed values, these effects were much smaller or negligible. Only older age, Black or African American racial identity, female biological sex, earlier variant period at first infection, non-vaccination status, and moderate self-rated mental or cognitive health before infection were associated with increased odds of long COVID. The demographic findings of this study are consistent with other large studies that have found higher prevalence of long COVID among people of older age [40] and female biological sex [39, 40, 42]. Our finding of greater likelihood of long COVID among Black or African Americans partially aligns with other analyses of odds differences by race [39, 41]. This interpretation should be considered cautiously, as we found considerable differences in distributions of race and education between this sample, all of whom joined the study prior to their first infection, and those who joined after their first infection.

This was the first study exploring the role of pre-infection functional status in a large dataset, and is strengthened by the use of both a valid and reliable survey [35] and clinician-documented EHR data sources [54]. We noted different unadjusted strengths and directions of associations from the self-reported versus EHR-derived functional status indicators (with moderate pre-infection self-rated mental health/cognition being associated with developing long COVID, while ability to perform physical daily activities was not). This was notable since many of the concepts contained in the “Finding of Functional Status and Activity” code hierarchy describe physical (e.g. motor and mobility) performance, with comparatively few cognitive or behavioral observations. Furthermore, in the adjusted odds, the EHR-derived diagnostic codes (pre-infection symptoms/conditions) for cognitive and mood disorders were also non-significant.

We propose several potential explanations for this discrepancy. First, it is likely that different latent constructs are measured by clinical observations encoded in the EHR compared to participant self-report on surveys. Self-report measures yield data on the respondent’s experience or perspective, whereas EHR indicators are likely derived from an external observer’s observations, often scored from structured tests or biometrics. The latter, as noted above, are predominant in this dataset’s “Finding of Functional Status and Activity” observation code, and are used in many other studies of functional status as an outcome of long COVID. However, physical functions are not the only contributor to functioning. In its International Classification of Functioning and Health (ICF), the WHO encodes functioning as the combined effect of body, context, and task factors on the ability to participate in daily life [55]. A study of pre-pandemic mental health and well-being indicators found similar discrepancies [30]. The concurrence between survey and EHR data in *All of Us* has also been found to vary widely for different diagnostic categories [56], so it is possible that there is a range of concurrence for functional status data in *All of Us* as well. Thus, self-report may provide data in this context that cannot be measured by clinical tools. Second, there is potential for under-detection of slight or moderate functional decreases in EHR data [57] particularly as functional status is rarely measured outside of acute or geriatric care contexts [58]. Third, systemic and payor barriers to obtaining mental health care services (and the resulting lack of diagnostic indications in the EHR) may have contributed to missingness of mental and cognitive health codes in the EHR. It is particularly important to consider differences between self-report and clinical observation data in persons with long COVID, as self-reported functional impairments in long COVID are sometimes dismissed in clinical settings [6] or measured with instruments that haven’t been validated with this population [59–61]. Exploring the concurrence of survey and EHR findings further is an excellent question for a future analysis. These findings reinforce the importance of collecting and using patient report alongside EHR data to provide comprehensive descriptions of pre- and post-illness risk and change [62].

This exploration of the pre-infection incidences of symptoms and conditions common in long COVID showed several insights. First, the distributions of these conditions relative to one another were remarkably similar between long COVID and recovered participants; with a few exceptions of slight magnitude, the conditions that were more prevalent in participants who later developed long COVID were also more prevalent in participants who later recovered. However, there was a striking difference in the *proportion* of each group experiencing each symptom, often twice as high or more in those who later developed post-infection long COVID symptoms.

Despite this difference in incidence, no single symptom contributed significantly to higher adjusted odds of long COVID; only sleep disorders approached significance. From a diagnostic perspective, this aligns with patient-led research findings that long COVID is a change in health status from pre-infection including both new and exacerbated symptoms and conditions compared to pre-COVID, and that symptom presentation can vary widely between individuals [5, 13, 22]. Many (but not all) participants who developed long COVID experienced pre-infection health issues found in long COVID phenotypes, with a wide range in the number of symptoms. Although the risk of long COVID was lower with no pre-infection symptoms, it was not eliminated.

The low correlations between most of these symptoms and weak groupings using first principal components suggest that, while some symptoms do appear more likely to co-occur, these do not suggest strong patterns of pre-infection symptom incidence in this sample. The exception was a moderate correlation between anxiety and depression, two of the most common [63] and frequently co-occurring [64] mental health conditions among U.S. adults, which may reflect the increasing prevalence of both disorders in recent years and during the pandemic in particular [65, 66]. It is important to note that, although the adjusted associations between prior symptoms and long COVID were non-significant (except at zero symptoms) in this study, the model’s variance inflation factors provide evidence that some symptoms’ odds ratios may have been underestimated. These results suggest that assessment of pre-infection health, including the presence of and interaction between ongoing health needs, should be considered in research and clinical management of long COVID, as previously effective management strategies may require adjustment. A future analysis could explore pre-versus post-infection changes in symptom and condition distribution, exacerbations versus new conditions, and dimensional reduction to explore and compare phenotypes.

The finding that earlier periods of first infection increased the likelihood of developing long COVID calls for additional context and exploration. Presumably, the opportunities to develop symptoms should have increased with time. This dataset’s cut-off date of July 2022 left little time for people in the last variant period to develop symptoms, and we did not control for time from first infection to first symptom. Additionally, we found that SARS-CoV-2 vaccination was associated with lower risk, consistent with other evidence from the same or similar time periods [27, 67]. However, we also found that the vaccination rate in this sample is lower than purpose-built COVID-19 cohort studies of the same period [27] which limited analysis. A future study could explore potential explanations for this such as differences in vaccination rates between regions or among the underrepresented groups from which *All of Us* purposefully over-samples, concurrence of drug encounters with self-reported vaccination (in the *All of Us* Minute Survey through early 2022), participant perspectives on vaccines (Minute Survey), or other systemic factors. Due to the relatively crude definition with which we were able to model these data, we could not determine the relationships between the date at first infection, the timing between vaccination and infection, the availability of vaccines starting in 2021, or the development of long COVID symptoms while retaining power to detect differences in risk by exposures of functional status and pre-infection symptom incidences, which were the primary focus of the present study. This relationship requires further examination beyond the scope of the present work. Growing evidence suggests that vaccination has complex period-cohort interactions affecting both infection and long COVID, potentially modified by number of doses, time since last dose (waning), infection timing relative to vaccination, boosting status, and biotechnological platform (e.g. mRNA, inactivated spike protein) [68–70]. (For an excellent summary of these challenges during the time period of this analysis, see Brannock et al, 2023 [27].) This important factor could be explored using a matched sub-sample of the present study’s sample, or in a future analysis of a later *All of Us* curated data repository in which more participants have had vaccination exposures and when the population has spent more time in COVID-19’s endemic phase.

### Limitations

This study has several limitations that impact interpretation. First, retrospective studies using EHR data are limited by detection bias [54, 57], a design aspect that could have affected this study in several ways. Several sources of relevant functional and disease severity data are unavailable in *All of Us*, and we were unsuccessful in finding proxies for severity of infection or long COVID symptoms. Detection bias may also have impacted classification of the outcome, particularly given our classification based on single incidences rather than persistence of symptoms. Second, the Recovered group may not be a complete representation. A challenge across population-based studies of COVID-19 is the difficulty verifying that participants without COVID-19 diagnostic and lab indicators were truly unexposed, particularly when using retrospective EHR data. It is possible that many participants who were not selected for this sample based on EHR evidence of COVID-19 infection had, in reality, been ill with the virus but were not identifiable as such because they did not report their illness or present for care, or experienced their first infection when the COPE survey was not administered so could not self-report their illness. Heterogeneity of health care delivery practices, documentation and measurement may have also reduced precision in the estimation of the primary predictor. These challenges are difficult to overcome without longitudinal patient report or other indices of symptom severity pre-versus post-infection, which is a limitation of the All of Us data compared to COVID-19 specific biobanks. Additionally, we were unable to estimate infection severity, an important factor in post-infection outcomes. Admitting diagnoses and narrative notes (where functional status and patient report of symptom severity are often described) are suppressed in *All of Us* for privacy, which precluded connections between incidences of COVID-19 and hospitalizations and the use of text-to-data methods of data extraction. Presciption of Remdesivir, used in acute treatment, was also explored as proxy for disease severity. The cut-off date for this *All of Us* data set predated FDA approval of Remdesivir by six months. About 800 participants had been prescribed remdesivir, presumably for experimental or off-label treatment. This relatively small number was insufficient to make inferences across the sample, and may have been confounded by access to clinical trials. Finally, we did not include several biomedical risk factors known to increase risk for long COVID such as cardiovascular disease, diabetes, and BMI. This was done to avoid over-parameterizing the model and retain power to detect differences in functional status as the primary predictor of interest. Due to their relationship with long COVID development, the biobehavioral relationships between these conditions and functional status should be more deeply explored in future investigations with these data.

Subsequent examinations of long COVID in these data could improve precision by combining collinear variables (e.g. using data reduction methods such as first principal components or hierarchical cluster analyses) or by weighting post-infection symptom occurrences if the symptom was also present prior to infection. Prospective studies using tools that measure disease and symptom severity, symptom or functional status change over time, and patient experiences, continue to be critical.

## Conclusions

In this model, neither pre-infection total incidences of long COVID symptoms (compared to the average of 4) nor pre-infection functional impairment increased risk of developing long COVID symptoms after infection. Long COVID was more likely for participants with self-reported moderate mental or cognitive health prior to infection, older age, female sex, Black racial identity, earlier variant at time of first infection, and non-vaccinated status. It was less likely for participants with no common long COVID symptoms occurring before infection. No single long COVID symptom prior to first infection affected the odds of long COVID after adjusting for demographics, disease factors, and functional status, however the impact of co-occurring symptoms requires further investigation. Functional status indicators exist in harmonized electronic health records data and have the potential to reveal the functional impacts of long COVID and improve diagnostic efficiency, but should be interpreted with caution and considered alongside self-report of function when available. Areas of future study include developing the diagnostic utility of functional status assessment, uncovering the relationship between long COVID-related functional changes and ageing, and tracking functional status changes from pre-infection to post-infection for people with post-COVID symptoms.

## Data Availability

These data are held in the publicly funded National Institutes of Health All of Us Research Program, Curated Data Repository Version 7.0 (CDR 7) Controlled tier C2022Q4R9. Summaries of these data with datetimes removed are available to the public in the Data Browser at https://databrowser.researchallofus.org/?_gl=1*6an21w*_ga*MTQ2NzA2OTQxNi4xNzUzOTA3MTk3*_ga_MQVR5DG2C4*czE3NTM5MDc1MjYkbzEkZzEkdDE3NTM5MDc1MzMkajUzJGwwJGg0ODU3NTY3Nzc. Per All of Us policy, researchers wishing to view these data in their entirety must apply and be approved for access, engage in yearly compliance training, and abide by a data user code of conduct. Details may be viewed at https://support.researchallofus.org/hc/en-us/articles/4415498292244-Overview-of-All-of-Us-Research-Program-Policies-for-Researchers. and at https://www.researchallofus.org/data-tools/data-access/?_gl=1*15sk6cc*_ga*MTQ2NzA2OTQxNi4xNzUzOTA3MTk3*_ga_MQVR5DG2C4*czE3NTM5MDc1MjYkbzEkZzEkdDE3NTM5MDc1NDIkajQ0JGwwJGg0ODU3NTY3Nzc.

## Acknowledgements

We extend gratitude to *All of Us* participants who generously agreed to share their ongoing health data. The first author also thanks the *All of Us* researcher workbench support staff for their expertise and service, the co-authors for their mentorship, Dr. Beth Fields for editing, Dr. Christine Sorkness for respiratory disease consultation, and Dr. Anoop Mayampurath and the University of Wisconsin Data Science Hub for dataset building support.

## Data Availability

This study used data from the *All of Us* Research Program’s Controlled Tier Dataset version 7, available to authorized users on the <u>Researcher Workbench</u> according to the *All of Us* <u>data access framework</u> and in compliance with the *<u>All of Us</u>* <u>data user code of conduct</u>. De-identified summary statistics with shifted date ranges are available for public browsing on the *<u>All</u> <u>of Us</u>* <u>Data Browser.</u> Registered and Controlled Tier data access information and registration is available at the All of Us Research Hub. Per the program’s Data User Agreement, the authors are not at liberty to share participant-level data.

## Supporting information

**S1 File. Appendices A-E.** Appendices with expanded description of cohort discovery; disease, symptom, and functional indicators; quantitative analysis of bias; model fitting notes; expanded results. All materials below are indexed in the S1 File.

**Table A.1. Identification of cohort - COVID-19 illness indicators.** All laboratory observations, COVID-19 Participant Experience (COPE) survey item responses, and diagnostic codes in SNOMED vocabulary indicating either SARS-CoV-2 infection or COVID-19 illness. Generated using queries for lab values, survey item responses, and diagnostic code incidences via the All of Us Researcher Workbench dataset builder.

**Table A.2. Long COVID symptoms/conditions used in Cohort discovery and classification.** Standard concept names, concept codes, source vocabularies, and included sub-concepts of long COVID symptoms queried via the *All of Us* Researcher Workbench dataset builder. Hierarchical relationships are as organized in the Athena relational database of the Observational Health Data Sciences and Informatics (OHDSI) and queried from the Observational Medical Outcomes Partnership – Common Data Model (OMOP – CDM) table structures.

**Table A.3. Vaccination Concepts.** Standard concept names, codes, vaccine type, and counts of vaccination data queried in *All of* Researcher Workbench dataset builder, Drug Encounters OMOP Table. Raw counts are from the unfiltered (N=104,993) sample of every vaccination encounter.

**Fig C.1. Distribution of propensity scores for pre-versus post-infection enrollment.**

Jitter point horizontal arrays showing the overlapping ranges in propensity for being in the long COVID group between the unmatched versus matched pre-infection enrollees (‘control’) and post-infection enrollees (‘treated’; excluded for the present study’s analysis). After matching pre- and post-enrollment participants, about a 45% overlap is seen between these two groups, indicating that participants may differ in one or more key demographic and disease aspects relating to when they enrolled in the study.

**Fig C.2. Standardized Mean Difference of demographic and disease characteristics by pre-versus post-infection enrollment.** Differences between standardized means of variant, race, sex, and education between participants who joined *All of Us* before versus after their first infection. Large differences are observed in race and education, with an overall distance of about 0.2 between these groups’ means. These demographic differences most likely reflect period differences in study enrollment efforts targeting under-represented groups in medical research, and may indicate that some of these groups are underrepresented in the present sample.

**Fig C.3. Distribution of propensity scores for 28-day versus 90-day symptom onset date.** Overlaps in propensity score between the participants classified as cases at 28 days (‘control’) and those at 90 days (‘treated’) is visualized in Fig C.3. There was no overlap between cases and controls by the 90-day alternative classification scheme (distance = 1.0).

**Fig C.4. Standardized Mean Difference of demographic and disease characteristics by 28-day versus 90-day symptom onset date.** Plot showing the difference in standardized mean propensity for classification as a long COVID case according to either a 28 day or a 90 day lag from first infection indicator. The matched samples show no significant difference in classification likelihood, indicating that, in this sample and study design, classifying participants as “cases” who had their first long COVID symptom 28 days or more after infection did not result in a significantly different case-versus-control distribution than classifying based on first symptom at 90 days or more.

**Table D.1. Variables for model.** Demographic, disease, pre-infection symptom, and pre-infection function variables (names, sources, and formats) used as covariates in regression models. Pre-infection variables are based on entry in the medical record between five years and four weeks prior to first infection date. Each variable’s intercept value or level is noted.

**Table D.2. Model summary.** The final model had 78 degrees of freedom from the fitted parameters. Model discrimination was good, with a maximum likelihood ratio X2 of 26194.58 and an area under the receiver operating curve (C) of 85%. The R2 – total variance explained by the fitted model – was 45%, indicating about half of the variance was unexplained by this model. Given the large sample size and the highly significant X2 of the maximum likelihood ratio (p <0.0001), this model was judged to have adequate fit for determining association.

**Fig D.1. Contributions of each variable to model effects.** Plot showing the contribution of each variable (X2) to the model’s omnibus effect size. Covariates are plotted on the Y axis; the least-significant contributor is at the top, and the rest are plotted down the axis in order of effect to the most-significant contributor at the bottom. The effect size is plotted on the X axis as X2 minus the degrees of freedom. Each variable’s effect is plotted as a dot intersecting the covariate with its effect size value. The top (smallest) contributor is the interaction between age and prior functional performance (X2=0.0, p=0.994); the bottom (largest) contributor is infection variant, (X2=5832.6, p<0.0000). **Table D.4. Variance inflation factor for each covariate.** Variance inflation factor (VIF) for the parameters of each covariate, an estimate of multicollinearity in the model. Variance inflation and collinearity can result from several covariates’ effects on the outcome increasing or decreasing at the same rate and/or magnitude (such that the effect of one could be predicted by or accounted for by the other). VIF was below 5.0 for most covariates, indicating low risk for multicollinearity. Seventeen parameter values (e.g. levels in a categorical variable or interaction) were above 5. None exceeded 10.

**Fig E.1. Time of first infection, by long COVID group.** Histogram of the number of participants with (darker blue) versus without (lighter blue) long COVID by first infection date through July 2022. The *Y* axis plots the number of participants ascending from zero (bottom) to over 1,500 (top); the *X* axis is a timeline from (left to right) January 1 2020 through July 31 2022. Both groups show a similar profile; they begin with a steep spike in infections about March 2020, a large and prolonged peak between April and September 2020, and briefer peaks between about October 2020 – January 2021 and in January 2022.

**Fig E.2. Among vaccinated participants, distributions of first infection and first vaccination date.** Two side-by-side bar graphs (controls on the left, cases on the right) of the timing of first infection and first full-series vaccination among the n=3,210 participants who were fully vaccinated. This graph suggests that, among participants with full vaccination, those with at least one long COVID symptom at any point had a higher disease incidence through early 2022 (spiking sharply in early 2020 and tapering gradually through 2021), and a higher uptake of both initial vaccination (December 2020 through about June 2021) and boosters (spiking in October 2021 and April 2022).

**Table E.1. Interaction: Age by functional status inferred from pre-infection EHR clinical observations.** Adjusted odds by age and pre-infection functional performance level (EHR code under “Finding of Functional Performance and Activity”) of developing long COVID.

**Table E.2. Interaction: Age by pre-infection self-reported physical ability.** Adjusted odds by age and self-rated ability to complete physical daily activities of developing long COVID.

**Table E.3. Interaction: Age by pre-infection total incident symptoms.** Adjusted odds by age and pre-infection symptom incidences of developing long COVID.

**Table E.4. Correlation matrix of pre-infection symptoms.** Correlation matrix of Pearson’s R2 correlation coefficient for all pre-infection incidences of long COVID symptoms included in the model.

## References

1. National Center for Health Statistics USCB. Household Pulse Survey - Long COVID 2024.

2. National Academies of Sciences Engineering and Medicine. A Long COVID Definition: A Chronic, Systemic Disease State with Profound Consequences. Washington, DC: The National Academies Press; 2024.

3. Cohen J, Rodgers YVM. Long COVID Prevalence, Disability, and Accommodations: Analysis Across Demographic Groups. J Occup Rehabil. 2024 Jun;34(2):335–49.

4. McCorkell L, G SA, H ED, Wei H, Akrami A. Patient-Led Research Collaborative: embedding patients in the Long COVID narrative. Pain Rep. 2021;6(1):e913.

5. Al-Aly Z, Davis H, McCorkell L, Soares L, Wulf-Hanson S, Iwasaki A, et al. Long COVID science, research and policy. Nat Med. 2024 Aug;30(8):2148–64.

6. Ladds E, Rushforth A, Wieringa S, Taylor S, Rayner C, Husain L, et al. Persistent symptoms after Covid-19: qualitative study of 114 “long Covid” patients and draft quality principles for services. BMC Health Serv Res. 2020 Dec 20;20(1):1144.

7. Mendelson M, Nel J, Blumberg L, Madhi SA, Dryden M, Stevens W, et al. Long-COVID: An evolving problem with an extensive impact. S Afr Med J. 2020 Nov 23;111(1):10–2.

8. The Lancet. Facing up to long COVID. Lancet. 2020 Dec 12;396(10266):1861.

9. Centers for Disease Control and Prevention. Long COVID or Post-COVID Conditions. 2023; Available from: https://www.cdc.gov/covid/long-term-effects/?CDC_AAref_Val= https://www.cdc.gov/coronavirus/2019-ncov/long-term-effects/.

10. Crook H, Raza S, Nowell J, Young M, Edison P. Long covid—mechanisms, risk factors, and management. BMJ. 2021 Jul 26;374:n1648.

11. Zhang H, Zang C, Xu Z, Zhang Y, Xu J, Bian J, et al. Data-driven identification of post-acute SARS-CoV-2 infection subphenotypes. Nature Medicine. 2023 Jan;29(1):226–35.

12. Davis EH, Assaf SG, Mccorkell L, Wei H, Low JR, Re’Em Y, et al. Characterizing long COVID in an international cohort: 7 months of symptoms and their impact. eClinicalMedicine. 2021 Aug;38:101019.

13. Davis H, Mccorkell L, Vogel J, Topol E. Long COVID: major findings, mechanisms and recommendations. Nature Reviews Microbiology. 2023 Mar;21(3):133–46.

14. Nielsen TB, Leth S, Pedersen M, Harbo HD, Nielsen CV, Laursen CH, et al. Mental Fatigue, Activities of Daily Living, Sick Leave and Functional Status among Patients with Long COVID: A Cross-Sectional Study. International journal of environmental research and public health. 2022 Nov 9;19(22).

15. Courtney MD, Edwards HE, Chang AM, Parker AW, Finlayson K, Bradbury C, et al. Improved functional ability and independence in activities of daily living for older adults at high risk of hospital readmission: a randomized controlled trial. J Eval Clin Pract. 2012 Feb;18(1):128–34.

16. Vanichkachorn G, Newcomb R, Cowl CT, Murad MH, Breeher L, Miller S, et al. Post-COVID-19 syndrome (long haul syndrome): Description of a multidisciplinary clinic at Mayo Clinic and characteristics of the initial patient cohort. Mayo Clin Proc. 2021 Jul;96(7):1782–91.

17. Gentilotti E, Górska A, Tami A, Gusinow R, Mirandola M, Baño RJ, et al. Clinical phenotypes and quality of life to define post-COVID-19 syndrome: a cluster analysis of the multinational, prospective ORCHESTRA cohort. eClinicalMedicine. 2023 Aug;62:102107.

18. Mina Y, Enose-Akahata Y, Hammoud DA, Videckis AJ, Narpala SR, O’Connell SE, et al. Deep Phenotyping of Neurologic Postacute Sequelae of SARS-CoV-2 Infection. Neurol Neuroimmunol Neuroinflamm. 2023 Jul;10(4).

19. Prabhakaran D, Day G, Munipalli B, Rush B, Pudalov L, Niazi S, et al. Neurophenotypes of COVID-19: risk factors and recovery trajectories. 20221221 ed2022.

20. Reese TJ, Blau H, Casiraghi E, Bergquist T, Loomba JJ, Callahan JT, et al. Generalisable long COVID subtypes: findings from the NIH N3C and RECOVER programmes. eBioMedicine. 2023 Jan;87:104413.

21. Al-Aly Z, Xie Y, Bowe B. High-dimensional characterization of post-acute sequelae of COVID-19. Nature. 2021;594(7862):259–64.

22. Thaweethai T, Jolley ES, Karlson WE, Levitan BE, Levy B, Mccomsey AG, et al. Development of a Definition of Postacute Sequelae of SARS-CoV-2 Infection. JAMA. 2023 Jun 13;329(22):1934.

23. Hou Y, Gu T, Ni Z, Shi X, Ranney ML, Mukherjee B. Global Prevalence of Long COVID, Its Subtypes, and Risk Factors: An Updated Systematic Review and Meta-analysis. Open Forum Infect Dis. 2025 Sep;12(9):ofaf533.

24. Callard F, Perego E. How and why patients made Long Covid. Soc Sci Med. 2021 Jan;268:113426.

25. Soriano JB, Murthy S, Marshall JC, Relan P, Diaz JV. A clinical case definition of post-COVID-19 condition by a Delphi consensus. Lancet Infect Dis. 2022 Apr;22(4):e102–e7.

26. Mandel HL, Colleen G, Abedian S, Ammar N, Bailey LC, Bennett TD, et al. Risk of post-acute sequelae of SARS-CoV-2 infection associated with pre-coronavirus disease obstructive sleep apnea diagnoses: an electronic health record-based analysis from the RECOVER initiative. Sleep. 2023 May 11.

27. Brannock MD, Chew RF, Preiss AJ, Hadley EC, Redfield S, McMurry JA, et al. Long COVID risk and pre-COVID vaccination in an EHR-based cohort study from the RECOVER program. Nat Commun. 2023 May 22;14(1):2914.

28. Hall JP, Kurth NK, McCorkell L, Goddard KS. Long COVID Among People With Preexisting Disabilities. Am J Public Health. 2024 Nov;114(11):1261–4.

29. Kuper H, Smythe T. Are people with disabilities at higher risk of COVID-19-related mortality?: a systematic review and meta-analysis. Public Health. 2023 Sep;222:115–24.

30. Zager Kocjan G, Verdnik J, Manfreda J, Komidar L, Lep Ž, Kobal Grum D, et al. Pre-pandemic lifestyle patterns and mental health outcomes among people reporting post-acute sequelae of COVID-19: evidence from a Slovenian population-based sample. BMC Public Health. 2025 Nov 21;25(1):4083.

31. National institutes of Health. All of Us, Curated data repository version 7, Controlled Tier C2022Q4R9. In: Health NIo, editor.: National Institutes of Health; 2022–2025.

32. Ramirez AH, Sulieman L, Schlueter DJ, Halvorson A, Qian J, Ratsimbazafy F, et al. The All of Us Research Program: Data quality, utility, and diversity. Patterns (N Y). 2022 Aug 12;3(8):100570.

33. Zeng C, Schlueter DJ, Tran TC, Babbar A, Cassini T, Bastarache LA, et al. Comparison of phenomic profiles in the All of Us Research Program against the US general population and the UK Biobank. J Am Med Inform Assoc. 2024 Apr 3;31(4):846–54.

34. 29 USC Ch. 28: Family and Medical Leave Act, (1993).

35. Hays RD, Schalet BD, Spritzer KL, Cella D. Two-item PROMIS® global physical and mental health scales. J Patient Rep Outcomes. 2017;1(1):2.

36. Centers for Disease Control and Prevention. New ICD-10-CM code for Post-COVID Conditions, following the 2019 Novel Coronavirus (COVID-19). In: Prevention CfDCa, editor.2021.

37. Markov PV, Ghafari M, Beer M, Lythgoe K, Simmonds P, Stilianakis NI, et al. The evolution of SARS-CoV-2. Nat Rev Microbiol. 2023 Jun;21(6):361–79.

38. National Healthcare Safety Network. COVID-19 Vaccination Modules: Key Terms: Centers for Disease Control and Prevention2025.

39. Jacobs MM, Evans E, Ellis C. Racial, ethnic, and sex disparities in the incidence and cognitive symptomology of long COVID-19. J Natl Med Assoc. 2023 Apr;115(2):233–43.

40. Vasilevskaya A, Mushtaque A, Tsang MY, Alwazan B, Herridge M, Cheung AM, et al. Sex and age affect acute and persisting COVID-19 illness. Sci Rep. 2023 Apr 13;13(1):6029.

41. Khullar D, Zhang Y, Zang C, Xu Z, Wang F, Weiner MG, et al. Racial/ethnic disparities in post-acute sequelae of SARS-CoV-2 infection in New York: an EHR-based cohort study from the RECOVER program. J Gen Intern Med. 2023 Apr;38(5):1127–36.

42. Shah DP, Thaweethai T, Karlson EW, Bonilla H, Horne BD, Mullington JM, et al. Sex Differences in Long COVID. JAMA Netw Open. 2025 Jan 2;8(1):e2455430.

43. Ford ND, Slaughter D, Edwards D, Dalton A, Perrine C, Vahratian A, et al. Long COVID and Significant Activity Limitation Among Adults, by Age — United States, June 1–13, 2022, to June 7–19, 2023. MMWR Morb Mortal Wkly Rep 2023. 2023;72:866–970.

44. Izaguirre P, Arakaki É, Boero JV, Zalazar Á, Ghirlanda M, Caruso D. Functional Status in Older Adults Following Hospitalization for Covid-19: A Cohort Study. Ann Geriatr Med Res. 2023 Sep 7.

45. U.S. Department of Health and Human Services: Office of Disease Prevention and Health Promotion. Healthy People 2030: Social Determinants of Health. 2025 [cited 2025 April 15]; Available from: https://odphp.health.gov/healthypeople/priority-areas/social-determinants-health/literature-summaries.

46. Case KR, Wang CP, Hosek MG, Lill SF, Howell AB, Taylor BS, et al. Health-related quality of life and social determinants of health following COVID-19 infection in a predominantly Latino population. J Patient Rep Outcomes. 2022 Jun 23;6(1):72.

47. Eligulashvili A, Darrell M, Gordon M, Jerome W, Fiori KP, Congdon S, et al. Patients with unmet social needs are at higher risks of developing severe long COVID-19 symptoms and neuropsychiatric sequela. Sci Rep. 2024 Apr 2;14(1):7743.

48. Centers for Medicare and Medicaid Services. Billing and Coding: Therapy Evaluation, Re-Evaluation and Formal Testing. A53309. Baltimore, MD: DHHS, CMS; 2020 [cited 2024 10/3/2024]; Available from: https://www.cms.gov/medicare-coverage-database/view/article.aspx?articleid=53309.

49. R Core Team. R: A language and environment for statistical computing. Vienna, Austria: R Foundation for Statistical Computing; 2022.

50. R Core Team and contributors worldwide. The R Stats Package. 4.3.0-4.4.3 ed. CRAN: R Foundation for Statistical Computing; 2025. p. R statistical functions.

51. Harrell FE. Regression Modeling Strategies. CRAN; 2024.

52. Wei T. Visualization of a Correlation Matrix (corrplot). 0.92 ed. CRAN: R Foundation for Statistical Computing; 2021.

53. Hadley Wickham MA, Jennifer Bryan, Winston Chang, Lucy D’Agostino McGowan, Romain François, Garrett Grolemund, Alex Hayes, Lionel Henry, Jim Hester, Max Kuhn, Thomas Lin Pedersen, Evan Miller, Stephan Milton Bache, Kirill Müller, Jeroen Ooms, David Robinson, Dana Paige Seidel, Vitalie Spinu, Kohske Takahashi, Davis Vaughan, Claus Wilke, Kara Woo, Hiroaki Yutani. Welcome to the tidyverse. 2.0.0 ed. CRAN: R Foundation for Statistical Computing; 2019.

54. Mandel HL, Shah SN, Bailey LC, Carton T, Chen Y, Esquenazi-Karonika S, et al. Opportunities and Challenges in Using Electronic Health Record Systems to Study Postacute Sequelae of SARS-CoV-2 Infection: Insights From the NIH RECOVER Initiative. J Med Internet Res. 2025 Mar 5;27:e59217.

55. World Health Organization. International Classification of Functioning, Disability and Health (ICF). Geneva: WHO2001.

56. Sulieman L, Cronin RM, Carroll RJ, Natarajan K, Marginean K, Mapes B, et al. Comparing medical history data derived from electronic health records and survey answers in the All of Us Research Program. J Am Med Inform Assoc. 2022 Jun 14;29(7):1131–41.

57. Deer RR, Rock AM, Vasilevsky N, Carmody L, Rando H, Anzalone JA, et al. Characterizing Long COVID: Deep Phenotype of a Complex Condition. eBioMedicine. 2021 Dec;74:103722.

58. Bogardus ST, Jr., Towle V, Williams CS, Desai MM, Inouye SK. What does the medical record reveal about functional status? A comparison of medical record and interview data. J Gen Intern Med. 2001 Nov;16(11):728–36.

59. Gutzeit J, Weiß M, Nürnberger C, Lemhöfer C, Appel KS, Pracht E, et al. Definitions and symptoms of the post-COVID syndrome: an updated systematic umbrella review. Eur Arch Psychiatry Clin Neurosci. 2024 Jul 25.

60. Lemhöfer C, Bahmer T, Baumbach P, Besteher B, Boekel A, Finke K, et al. Variations and Predictors of Post-COVID Syndrome Severity in Patients Attending a Post-COVID Outpatient Clinic. J Clin Med. 2023 Jun 12;12(12).

61. Baalmann AK, Blome C, Stoletzki N, Donhauser T, Apfelbacher C, Piontek K. Patient-reported outcome measures for post-COVID-19 condition: a systematic review of instruments and measurement properties. BMJ Open. 2024 Dec 20;14(12):e084202.

62. Brown JA, Lee AJ, Fernhoff K, Pistone T, Pasquini L, Wise AB, et al. Functional network collapse in neurodegenerative disease. Nat Commun. 2025 Nov 21;16(1):10273.

63. Terlizzi EP, Zablotsky B. Symptoms of anxiety and depression among adults: United States, 2019 and 2022. Hyattsville, MD: National Center for Health Statistics2024.

64. Hopwood M. Anxiety Symptoms in Patients with Major Depressive Disorder: Commentary on Prevalence and Clinical Implications. Neurol Ther. 2023 Apr;12(Suppl 1):5–12.

65. Pathak Y, Makk-Frid E. Population Estimates of Self-Reported Depression and Anxiety in the US From a National Survey: Cross-Sectional Survey Study. Interact J Med Res. 2025 Apr 16;14:e70626.

66. The Lancet. Global prevalence and burden of depressive and anxiety disorders in 204 countries and territories in 2020 due to the COVID-19 pandemic. Lancet. 2021 Nov 6;398(10312):1700–12.

67. Sinclair JE, Mayfield HJ, Lu H, Brown SJ, Moghaddam T, Waller M, et al. Estimating risk of long COVID using a Bayesian network-based decision support tool. Vaccine. 2025 Dec 18;72:128127.

68. Al-Aly Z, Bowe B, Xie Y. Long COVID after breakthrough SARS-CoV-2 infection. Nat Med. 2022 Jul;28(7):1461–7.

69. Cai M, Xie Y, Al-Aly Z. Association of 2024-2025 Covid-19 Vaccine with Covid-19 Outcomes in U.S. Veterans. N Engl J Med. 2025 Oct 23;393(16):1612–23.

70. Guimarães GN, Brunetti NS, De Lima DG, Proenca-Modena JL, Farias AS. Vaccination and COVID-19: impact on long-COVID. Front Immunol. 2025;16:1686572.

