## Appendices A-E for "Long COVID risk by pre-infection symptoms and functional status: A retrospective cohort study of data from the *All of Us* Research Program"

### Supplemental: Appendices

| Section | Title | Page no. |
|---|---|---|
| Appendix A | Long COVID symptoms discovery and analysis | 2 |
| Appendix B | Functional status indicators | 13 |
| Appendix C | Sensitivity analysis of cohort discovery | 16 |
| Appendix D | Model specifications and statistics | 22 |
| Appendix E | Expanded results | 33 |
| References | | 38 |

### Appendix A. Long COVID symptoms discovery and analysis

#### All of Us Research Program data structure

All EMR observations in All of Us are harmonized in format of the Athena relational database of the Observational Health Data Sciences and Informatics (OHDSI), and structured according to the Observational Medical Outcomes Partnership – Common Data Model OMOP-CDM; 1,2. This ensures consistency across data collection sites and regions, and comparability with billing and treatment codes. All of Us utilizes community-relevant recruitment methods to over-recruit from geographic and demographic groups that are under-represented in medical and health research. The program, data snapshots, and data methods can be viewed at https://www.researchallofus.org/data-tools/.

Identification of cohort using COVID-19 illness indicators

Table S.A.1 Shows the variables and responses used to identify All of Us participants with at least one COVID-19 infection for inclusion in this study. Laboratory observations with positive test indications, responses on any version of the COVID-19 Participant Experience (COPE) survey, and COVID-19 related ICD-10 diagnostic codes were queried using the All of Us Researcher Workbench concept set builder, which was then applied as the inclusion filter.

##### Table A.1. Identification of cohort - COVID-19 illness indicators

| Name | Concept ID | Vocab | Code | Roll-up Count* | Item Count* |
|---|---|---|---|---|---|
| Laboratory observations | | | | | |
| 2019-ncov coronavirus, sars-cov-2/2019-ncov (covid-19), any technique, multiple types or subtypes (includes all targets), non-cdc | 40218804 | HCPCS | U0002 | 0 | 1,020 |
| 2019-nCoV Coronavirus, SARS-CoV-2/2019-nCoV (COVID-19), any technique, multiple types or subtypes (includes all targets), non-CDC, making use of high throughput technologies as described by CMS-2020-01-R | 704058 | HCPCS | U0004 | 0 | 1 |
| Infectious agent detection by nucleic acid (DNA or RNA); severe acute respiratory syndrome coronavirus 2 (SARS-CoV-2) (Coronavirus disease [COVID-19]), amplified probe technique, making use of high throughput technologies as described by CMS-2020-01-R | 704059 | HCPCS | U0003 | 0 | 1,143 |
| Influenza virus A and B and SARS-CoV-2 (COVID-19) and SARS-related CoV RNA panel | 36660845 | LOINC | LP418968-6 | 985 | 0 |
| Influenza virus A and B and SARS-CoV-2 (COVID-19) and SARS-related CoV RNA panel - Respiratory specimen by NAA with probe detection | 36661384 | LOINC | 95380-2 | 0 | 985 |
| Influenza virus A and B and SARS-CoV-2 (COVID-19) and SARS-related CoV RNA panel \| Respiratory specimen \| Microbiology Panels | 36661218 | LOINC | LP419290-4 | 985 | 0 |

| | | | | | |
|---|---|---|---|---|---|
| Influenza virus A and B and SARS-CoV-2 (COVID-19) RNA panel - Respiratory specimen by NAA with probe detection | 36661376 | LOINC | 95422-2 | 0 | 330 |
| Measurement of Severe acute respiratory syndrome coronavirus 2 (SARS-CoV-2) | 756055 | OMOP Extension | OMOP4873969 | 0 | 384 |
| Measurement of Severe acute respiratory syndrome coronavirus 2 antibody | 37310258 | SNOMED | 1.24046E+15 | 0 | 263 |
| SARS-CoV-2 (COVID-19) | 36662140 | LOINC | LP417540-4 | 92,959 | 0 |
| SARS-CoV-2 (COVID-19) Ab | 36661733 | LOINC | LP417914-1 | 9,362 | 0 |
| SARS-CoV-2 (COVID-19) Ab [Interpretation] in Serum or Plasma | 723480 | LOINC | 94661-6 | 0 | 3,037 |
| SARS-CoV-2 (COVID-19) Ab [Presence] in Serum or Plasma by Immunoassay | 586515 | LOINC | 94762-2 | 0 | 1,146 |
| SARS-CoV-2 (COVID-19) Ab [Units/volume] in Serum or Plasma by Immunoassay | 586522 | LOINC | 94769-7 | 0 | 309 |
| SARS-CoV-2 (COVID-19) Ab \| Serum or Plasma \| Microbiology | 36661221 | LOINC | LP418684-9 | 4,130 | 0 |
| SARS-CoV-2 (COVID-19) Ab panel | 36661883 | LOINC | LP418122-0 | 2 | 0 |
| SARS-CoV-2 (COVID-19) Ab panel - Serum or Plasma by Immunoassay | 706179 | LOINC | 94504-8 | 0 | 2 |
| SARS-CoV-2 (COVID-19) Ab panel \| Serum or Plasma \| Microbiology Panels | 36661105 | LOINC | LP419286-2 | 2 | 0 |
| SARS-CoV-2 (COVID-19) Ag | 36661764 | LOINC | LP418019-8 | 1,262 | 0 |
| SARS-CoV-2 (COVID-19) Ag [Presence] in Respiratory specimen by Rapid immunoassay | 723477 | LOINC | 94558-4 | 0 | 1,222 |
| SARS-CoV-2 (COVID-19) Ag [Presence] in Upper respiratory specimen by Immunoassay | 36032419 | LOINC | 96119-3 | 0 | 40 |
| SARS-CoV-2 (COVID-19) Ag \| Respiratory specimen \| Microbiology | 36660801 | LOINC | LP418693-0 | 1,222 | 0 |
| SARS-CoV-2 (COVID-19) Ag \| Upper respiratory specimen \| Microbiology | 36033457 | LOINC | LP420931-0 | 40 | 0 |
| SARS-CoV-2 (COVID-19) clade | 1620066 | LOINC | LP422736-1 | 47 | 0 |
| SARS-CoV-2 (COVID-19) clade [Type] in Specimen by Molecular genetics method | 36033653 | LOINC | 96896-6 | 0 | 47 |
| SARS-CoV-2 (COVID-19) clade \| XXX \| Microbiology | 1618285 | LOINC | LP427406-6 | 47 | 0 |
| SARS-CoV-2 (COVID-19) IgA | 36662109 | LOINC | LP418430-7 | 167 | 0 |
| SARS-CoV-2 (COVID-19) IgA \| Serum or Plasma \| Microbiology | 36660931 | LOINC | LP418685-6 | 167 | 0 |

| | | | | | |
|---|---|---|---|---|---|
| SARS-CoV-2 (COVID-19) IgA Ab [Presence] in Serum or Plasma by Immunoassay | 723473 | LOINC | 94562-6 | 0 | 167 |
| SARS-CoV-2 (COVID-19) IgG | 36661886 | LOINC | LP417915-8 | 5,025 | 0 |
| SARS-CoV-2 (COVID-19) IgG \| Serum or Plasma \| Microbiology | 36661046 | LOINC | LP418688-0 | 3,500 | 0 |
| SARS-CoV-2 (COVID-19) IgG \| Serum, Plasma or Blood \| Microbiology | 36660768 | LOINC | LP418689-8 | 2,090 | 0 |
| SARS-CoV-2 (COVID-19) IgG Ab [Presence] in Serum or Plasma by Immunoassay | 723474 | LOINC | 94563-4 | 0 | 2,920 |
| SARS-CoV-2 (COVID-19) IgG Ab [Presence] in Serum, Plasma or Blood by Rapid immunoassay | 706181 | LOINC | 94507-1 | 0 | 2,090 |
| SARS-CoV-2 (COVID-19) IgG Ab [Units/volume] in Serum or Plasma by Immunoassay | 706177 | LOINC | 94505-5 | 0 | 749 |
| SARS-CoV-2 (COVID-19) IgG+IgM | 36661646 | LOINC | LP417956-2 | 991 | 0 |
| SARS-CoV-2 (COVID-19) IgG+IgM \| Serum or Plasma \| Microbiology | 36660914 | LOINC | LP418690-6 | 991 | 0 |
| SARS-CoV-2 (COVID-19) IgG+IgM Ab [Presence] in Serum or Plasma by Immunoassay | 723479 | LOINC | 94547-7 | 0 | 991 |
| SARS-CoV-2 (COVID-19) IgM | 36661975 | LOINC | LP417916-6 | 396 | 0 |
| SARS-CoV-2 (COVID-19) IgM \| Serum or Plasma \| Microbiology | 36661274 | LOINC | LP418691-4 | 299 | 0 |
| SARS-CoV-2 (COVID-19) IgM \| Serum, Plasma or Blood \| Microbiology | 36660777 | LOINC | LP418692-2 | 97 | 0 |
| SARS-CoV-2 (COVID-19) IgM Ab [Presence] in Serum or Plasma by Immunoassay | 723475 | LOINC | 94564-2 | 0 | 275 |
| SARS-CoV-2 (COVID-19) IgM Ab [Presence] in Serum, Plasma or Blood by Rapid immunoassay | 706180 | LOINC | 94508-9 | 0 | 97 |
| SARS-CoV-2 (COVID-19) IgM Ab [Units/volume] in Serum or Plasma by Immunoassay | 706178 | LOINC | 94506-3 | 0 | 37 |
| SARS-CoV-2 (COVID-19) lineage | 1619966 | LOINC | LP422739-5 | 17 | 0 |
| SARS-CoV-2 (COVID-19) lineage [Identifier] in Specimen by Molecular genetics method | 36033652 | LOINC | 96895-8 | 0 | 17 |
| SARS-CoV-2 (COVID-19) lineage \| XXX \| Microbiology | 1618914 | LOINC | LP427405-8 | 17 | 0 |
| SARS-CoV-2 (COVID-19) N gene | 36661396 | LOINC | LP417599-0 | 1,151 | 0 |
| SARS-CoV-2 (COVID-19) N gene [Presence] in Nasopharynx by NAA with probe detection | 715272 | LOINC | 94760-6 | 0 | 36 |
| SARS-CoV-2 (COVID-19) N gene [Presence] in Nose by NAA with probe detection | 757678 | LOINC | 95409-9 | 0 | 9 |

| | | | | | |
|---|---|---|---|---|---|
| SARS-CoV-2 (COVID-19) N gene [Presence] in Respiratory specimen by NAA with probe detection | 706161 | LOINC | 94533-7 | 0 | 671 |
| SARS-CoV-2 (COVID-19) N gene [Presence] in Respiratory specimen by Nucleic acid amplification using CDC primer-probe set N1 | 586524 | LOINC | 94756-4 | 0 | 468 |
| SARS-CoV-2 (COVID-19) N gene \| Nasopharynx \| Microbiology | 36660752 | LOINC | LP418702-9 | 36 | 0 |
| SARS-CoV-2 (COVID-19) N gene \| Nose \| Microbiology | 36660970 | LOINC | LP419179-9 | 9 | 0 |
| SARS-CoV-2 (COVID-19) N gene \| Respiratory specimen \| Microbiology | 36661286 | LOINC | LP418703-7 | 1,107 | 0 |
| SARS-CoV-2 (COVID-19) ORF1ab region | 36661401 | LOINC | LP417906-7 | 1,267 | 0 |
| SARS-CoV-2 (COVID-19) ORF1ab region [Presence] in Respiratory specimen by NAA with probe detection | 723478 | LOINC | 94559-2 | 0 | 1,265 |
| SARS-CoV-2 (COVID-19) ORF1ab region [Presence] in Specimen by NAA with probe detection | 723464 | LOINC | 94639-2 | 0 | 2 |
| SARS-CoV-2 (COVID-19) ORF1ab region \| Respiratory specimen \| Microbiology | 36661250 | LOINC | LP418706-0 | 1,265 | 0 |
| SARS-CoV-2 (COVID-19) ORF1ab region \| XXX \| Microbiology | 36661194 | LOINC | LP418707-8 | 2 | 0 |
| SARS-CoV-2 (COVID-19) RdRp gene | 36661801 | LOINC | LP417598-2 | 3,774 | 0 |
| SARS-CoV-2 (COVID-19) RdRp gene [Presence] in Respiratory specimen by NAA with probe detection | 706160 | LOINC | 94534-5 | 0 | 3,764 |
| SARS-CoV-2 (COVID-19) RdRp gene [Presence] in Specimen by NAA with probe detection | 706173 | LOINC | 94314-2 | 0 | 18 |
| SARS-CoV-2 (COVID-19) RdRp gene \| Respiratory specimen \| Microbiology | 36660902 | LOINC | LP418708-6 | 3,764 | 0 |
| SARS-CoV-2 (COVID-19) RdRp gene \| XXX \| Microbiology | 36660887 | LOINC | LP418709-4 | 18 | 0 |
| SARS-CoV-2 (COVID-19) RNA | 36661507 | LOINC | LP417541-2 | 89,445 | 0 |
| SARS-CoV-2 (COVID-19) RNA [Cycle Threshold #] in Respiratory specimen by NAA with probe detection | 586528 | LOINC | 94745-7 | 0 | 1 |
| SARS-CoV-2 (COVID-19) RNA [Presence] in Nasopharynx by NAA with non-probe detection | 723476 | LOINC | 94565-9 | 0 | 2,005 |
| SARS-CoV-2 (COVID-19) RNA [Presence] in Nasopharynx by NAA with probe detection | 586526 | LOINC | 94759-8 | 0 | 273 |
| SARS-CoV-2 (COVID-19) RNA [Presence] in Respiratory specimen by NAA with probe detection | 706163 | LOINC | 94500-6 | 0 | 63,160 |

| | | | | | |
|---|---|---|---|---|---|
| SARS-CoV-2 (COVID-19) RNA [Presence] in Saliva (oral fluid) by Sequencing | 715261 | LOINC | 94822-4 | 0 | 107 |
| SARS-CoV-2 (COVID-19) RNA [Presence] in Specimen by NAA with probe detection | 706170 | LOINC | 94309-2 | 0 | 27,394 |
| SARS-CoV-2 (COVID-19) RNA \| Nasopharynx \| Microbiology | 36661317 | LOINC | LP418694-8 | 2,274 | 0 |
| SARS-CoV-2 (COVID-19) RNA \| Respiratory specimen \| Microbiology | 36661115 | LOINC | LP418695-5 | 63,160 | 0 |
| SARS-CoV-2 (COVID-19) RNA \| Saliva \| Microbiology | 36660966 | LOINC | LP418696-3 | 107 | 0 |
| SARS-CoV-2 (COVID-19) RNA \| XXX \| Microbiology | 36661244 | LOINC | LP418698-9 | 27,394 | 0 |
| SARS-CoV-2 (COVID-19) RNA panel | 36661522 | LOINC | LP417539-6 | 5,080 | 0 |
| SARS-CoV-2 (COVID-19) RNA panel - Respiratory specimen by NAA with probe detection | 706158 | LOINC | 94531-1 | 0 | 1,342 |
| SARS-CoV-2 (COVID-19) RNA panel - Specimen by NAA with probe detection | 706169 | LOINC | 94306-8 | 0 | 3,740 |
| SARS-CoV-2 (COVID-19) RNA panel \| Respiratory specimen \| Microbiology Panels | 36661036 | LOINC | LP419288-8 | 1,342 | 0 |
| SARS-CoV-2 (COVID-19) RNA panel \| XXX \| Microbiology Panels | 36660924 | LOINC | LP419289-6 | 3,740 | 0 |
| SARS-CoV-2 (COVID-19) S protein RBD neutralizing antibody [Presence] in Serum or Plasma by sVNT | 36031734 | LOINC | 96603-6 | 0 | 476 |
| SARS-CoV-2 (COVID-19) sequencing and identification panel | 1620099 | LOINC | LP422740-3 | 40 | 0 |
| SARS-CoV-2 (COVID-19) sequencing and identification panel - Specimen by Molecular genetics method | 36033651 | LOINC | 96894-1 | 0 | 40 |
| SARS-CoV-2 (COVID-19) sequencing and identification panel \| XXX \| Microbiology Panels | 1618441 | LOINC | LP427524-6 | 40 | 0 |
| SARS-CoV-2 (COVID-19) spike protein receptor binding domain (RBD) | 36033856 | LOINC | LP421235-5 | 476 | 0 |
| SARS-CoV-2 (COVID-19) spike protein receptor binding domain (RBD) neutralizing antibody | 36033858 | LOINC | LP421234-8 | 476 | 0 |
| SARS-CoV-2 (COVID-19) spike protein receptor binding domain (RBD) neutralizing antibody \| Serum or Plasma \| Microbiology | 36033625 | LOINC | LP421840-2 | 476 | 0 |
| SARS-CoV+SARS-CoV-2 (COVID-19) | 36661687 | LOINC | LP418774-8 | 1,075 | 0 |
| SARS-CoV+SARS-CoV-2 (COVID-19) Ag | 36661520 | LOINC | LP418762-3 | 1,075 | 0 |
| SARS-CoV+SARS-CoV-2 (COVID-19) Ag [Presence] in Respiratory specimen by Rapid immunoassay | 757685 | LOINC | 95209-3 | 0 | 1,075 |

| | | | | | |
|---|---|---|---|---|---|
| SARS-CoV+SARS-CoV-2 (COVID-19) Ag \| Respiratory specimen \| Microbiology | 1618075 | LOINC | LP427654-1 | 1,075 | 0 |
| COVID-19 Participant Experience (COPE) survey response | | | | | |
| COPE Survey (Any version): In the past month, have you been sick for more than one day with a new illness related to COVID-19 or flu-like symptoms?: Yes | 1332898 | | | 9,138 | |
| Diagnostic codes | | | | | |
| COVID-19 | 37311061 | SNOMED | 840539006 | 17,384 | 17,384 |
| Lower respiratory infection caused by SARS-CoV-2 | 3663281 | SNOMED | 8.8053E+17 | 1,663 | 0 |
| * = Among all program participants | | | | | |

Table Caption: All laboratory observations, COVID-19 Participant Experience (COPE) survey item responses, and diagnostic codes in SNOMED vocabulary indicating either SARS-CoV-2 infection or COVID-19 illness. Generated using queries for lab values, survey item responses, and diagnostic code incidences via the All of Us Researcher Workbench dataset builder.

#### Symptom-based classification of long COVID groups: Long COVID (case) vs. Recovered (control) groups

An initial list of sequelae was compiled from phenotyping studies e.g., 3,4. This list was then cross-checked with sequelae listed by the CDC,5 the CDC's ICD-10-CM emergency billing code for "Post COVID condition, Unspecified" U07.7)6, standard concept codes referring to diagnosis and symptom concepts found in the All of Us based reproduction of the RECOVER study phenotyping approach as shared in a featured All of Us workspace7 and co-author knowledge of patient experience encountered in a long COVID follow-up clinic. These were collapsed into diagnostically-related groups as organized under the Athena Health Observational Medical Outcomes Partnership – Common Data Model (OMOP-CDM) harmonized data vocabulary 1,2. (Athena is a hierarchical compendium of health concepts (e.g. diagnoses, observations, medications) organized into tree-like branching relationships. Distinct "descendent" concepts branch off of broad categories of "ancestor" concepts.) The final list consisted of 44 symptom/sequelae ancestor categories.

Using the All of Us Data Browser and Researcher Workbench, each symptom concept code was queried singly to create a concept set incorporating all occurrences of all descendent codes contained within that symptom for each participant with at least one COVID-19 infection. These were then arranged by date of recording. The data were then reduced to two occurrences; the first occurrence between 28 days and 5 years prior to the first COVID infection (pre-infection incidence) and the second occurrence being the first code 28 days or more after the first COVID infection (post-infection incidence, marking a participant as a long COVID group member). This was performed iteratively for each long COVID symptom/condition dataset individually, creating one dataset for each symptom containing only the first pre- and post-infection (if any) incidences for each participant. These datasets were then joined one by one in alphabetical order to the main dataset, using person ID number as the foreign key. The list of queried diagnostic categories is in Table S.A.2. Under these categories were 1,013 discrete descendent diagnostic concepts. We chose not to remove descendent concepts containing attributions to specific (non-COVID-19) diseases (e.g., "cancer-related fatigue", "memory impairment due to multiple sclerosis"), resulting in an attribution-agnostic grouping. This was for several reasons. First and most broadly, there is probable variation in how post-COVID pathophysiology interacts with other diseases processes; therefore the possibility of development or worsening of these health outcomes in the setting of other premorbid diagnoses could not be assumed to be independent of the physical toll of COVID recovery, and some outcomes may have been potentiated by this toll. Second, the increased granularity of having all types of a given diagnosis provided opportunities for future analysis that would not be possible had we removed or collapsed their diagnostic variations. Third and finally, we found the patient counts of many diagnostically-attributed outcomes were frequently small (fewer than n=50), necessitating collapse into other categories for statistical analysis and All of Us reporting rules, and exerting little or negligible pull on the symptoms' effect given the size of this dataset.

Table A.2. Long COVID symptoms/conditions used in Cohort discovery and classification

| Standard Concepts | OMOP Concept ID | Source | Vocab | Code | Strings collapsed |
|---|---|---|---|---|---|
| Abdominal pain | 200219 | Standard | SNOMED | 21522001 | "abdominal" |
| Anxiety | 441542 | Standard | SNOMED | 48694002 | "anxiety", "anxi*" |
| Chest pain | 77670 | Standard | SNOMED | 29857009 | "chest", "cardiac" |
| Chronic fatigue syndrome | 432738 | Standard | SNOMED | 52702003 | "chronic fatigue", "myalgic encephalomyelitis" |
| Cognitive disorder (collapsed into “impaired cognition”) | 40480615 | Standard | SNOMED | 443265004 | "cognitive", "neurocognitive" |
| Cognitive function finding (collapsed into “impaired cognition”) | 4162723 | Standard | SNOMED | 373930000 | [not collapsed, heterogeneous] |
| Cough | 254761 | Standard | SNOMED | 49727002 | "cough", "clearing throat" |
| Depression screening positive | 762504 | Standard | SNOMED | 4.28181E+14 | "depression", "screening" |
| Depressive disorder | 440383 | Standard | SNOMED | 35489007 | "depressive", "depression" |
| Depressive episode | 3656234 | Standard | SNOMED | 871840004 | |
| Diarrhea | 196523 | Standard | SNOMED | 62315008 | "diarrhea", "diarrheal" |
| Disorder of menstruation | 443431 | Standard | SNOMED | 386804004 | "*men* |
| Disturbance in sleep behavior (collapsed into “Sleep”) | 4204989 | Standard | SNOMED | 53888004 | "sleep" |
| Dizziness | 4223938 | Standard | SNOMED | 404640003 | "dizziness", "dizzy", "vertigo" |
| Dyspnea | 312437 | Standard | SNOMED | 267036007 | "dyspnea", "*pnea", "gasping" |
| Eruption | 140214 | Standard | SNOMED | 271807003 | "eruption", "acne*", "rash", "eruption", "erythroderma", "exanthematous", "Fox-Fordyce", "rocacea", "Keratin*", "pityriasis", "psoriasis", "psoriatic", "dermatitis" |
| Fatigue | 4223659 | Standard | SNOMED | 84229001 | "asthenia" |
| Fever | 437663 | Standard | SNOMED | 386661006 | "fever", "*pyrexia" |
| Finding of pattern of menstrual cycle (collapsed into “menstrual disorder”) | 4095940 | Standard | SNOMED | 248968007 | "menstru*", "amenorrhea" |
| Finding of sexual function | 4041277 | Standard | SNOMED | 118202007 | |
| Headache | 378253 | Standard | SNOMED | 25064002 | "headache", "head" |

| | | | | | |
|---|---|---|---|---|---|
| Impaired Cognition | 443432 | Standard | SNOMED | 386806002 | "cognitive", "impairment", "behavioral" |
| Irregular periods (collapsed into "menstrual disorder") | 196168 | Standard | SNOMED | 80182007 | |
| Joint pain | 77074 | Standard | SNOMED | 57676002 | "arthralgia", "joint" |
| Lightheadedness | 4297376 | Standard | SNOMED | 386705008 | "malaise and fatigue", "fatigue". Collapsed into "Fatigue" |
| Loss of sense of smell | 4185711 | Standard | SNOMED | 44169009 | |
| Loss of taste | 4289517 | Standard | SNOMED | 36955009 | |
| Malaise (under "Lightheadedness" heirarchy) | 4272240 | Standard | SNOMED | 367391008 | |
| Muscle fatigue | 4214612 | Standard | SNOMED | 80449002 | "muscle" |
| Muscle pain | 442752 | Standard | SNOMED | 68962001 | "myalgia*", "pain", "muscle", "pleurodynia", "fibrositis", "fibromyalgia", "polymyalgia", "claudication", "migraine" |
| Musculoskeletal chest pain | 4092930 | Standard | SNOMED | 281245003 | "pleurodynia", "myalgia", "pain", "Scapulalgia", "Xiphodynia", "syndrome" |
| Palpitations | 315078 | Standard | SNOMED | 80313002 | "heart", "palpitations" |
| Paraesthesia | 4236484 | Standard | SNOMED | 91019004 | ""*esthesia", "sensation", "pins", |
| Post-acute COVID-19 | 705076 | Standard | OMOP Extension | OMOP5160861 | U09.9 |
| Postural orthostatic tachycardia syndrome (POTS) | 4159659 | | SNOMED | 371073003 | (No hierarchy) |
| Postviral fatigue syndrome | 4202045 | Standard | SNOMED | 51771007 | |
| Sleep disorder (collapsed into "Sleep") | 435524 | Standard | SNOMED | 39898005 | "sleep*", "dream", "*somnia", "somnolence" |
| Tachycardia | 444070 | Standard | SNOMED | 3424008 | "tachycardia*" |

Table caption: Standard concept names, concept codes, source vocabularies, and included sub-concepts of long COVID symptoms queried via the *All of Us* Researcher Workbench dataset builder. Hierarchical relationships are as organized in the Athena relational database of the Observational Health Data Sciences and Informatics (OHDSI) and queried from the Observational Medical Outcomes Partnership – Common Data Model (OMOP – CDM) table structures.

#### Concept filters of non-long COVID related diagnostic concepts

After importing every incidence of the condition codes, concepts were removed that were not applicable to long COVID diagnosis, or were redundant (covered under other concept hierarchies). Under the “Cognition” and “Impaired Cognition” concept, “Normal Cognition” was recoded to “0” to match the presumptive absence of impairment in other participants with no data value (N/A). Psychological concepts from the DSM series were also removed. Filters were run as follows:

```
filter(!(standard_concept_name %in% c('Normal cognition',
                                      'Suicidal behavior',
                                      'Delusion of persecution',
                                      'Homicidal thoughts',
                                      'Worried',
                                      'Anxiety about body function or health',
                                      'Suicidal intent',
                                      'Delusions',
                                      'Thoughts of violence',
                                      'Deficient knowledge of preconception health
    practices',
                                      'Cognitive developmental delay',
                                      'Below average intellect',
                                      'Paranoid delusion',
                                      'Anxiety about treatment',
                                      'Thoughts of self harm',
                                      'Has access to planned means of suicide',
                                      'Repetitive routines',
                                      'Obsessional thoughts',
                                      'Planning suicide',
                                      'Dangerous and harmful thoughts',
                                      'Low intelligence',
                                      'Grinding teeth',
                                      'Human immunodeficiency virus infection with
    cognitive impairment', #redundant with other codes
                                      'Suicidal',
                                      'Suicidal thoughts',
                                      'Mood-congruent delusion',
                                      'Paranoid ideation'
                                     )))
```

```
Cognition %>%
    filter(standard_concept_name != "Attention deficit hyperactivity disorder" &
             standard_concept_name != "Attention deficit hyperactivity disorder,
    predominantly inattentive type" &
             standard_concept_name != "Child attention deficit disorder" &
             standard_concept_name != "Undifferentiated attention deficit disorder" &
             standard_concept_name != "Attention deficit hyperactivity disorder,
    predominantly hyperactive impulsive type" &
           standard_concept_name != "Attention deficit hyperactivity disorder, combined
    type" &
          standard_concept_name != "Hyperkinetic conduct disorder" &
          standard_concept_name != "Developmental coordination disorder")
```

Under the “Finding of Functional Performance and Activity” concept, psychological concepts from the DSM series and developmental diagnoses were removed as follows:

```
functional_performance %>%
    filter(standard_concept_name != "Attention deficit hyperactivity disorder" &
            standard_concept_name != "Attention deficit hyperactivity disorder,
    predominantly inattentive type" &
            standard_concept_name != "Child attention deficit disorder" &
            standard_concept_name != "Undifferentiated attention deficit disorder" &
            standard_concept_name != "Attention deficit hyperactivity disorder,
    predominantly hyperactive impulsive type" &
           standard_concept_name != "Attention deficit hyperactivity disorder, combined
    type" &
          standard_concept_name != "Hyperkinetic conduct disorder" &
          standard_concept_name != "Developmental coordination disorder" &
          standard_concept_name != "Developmental disorder of motor function" &
          standard_concept_name != "Developmental delay in fine motor function" &
          standard_concept_name != "Hyperkinesis with developmental delay" &
          standard_concept_name != "Ineffective breathing pattern" &
          standard_concept_name != "Adjustment to life threatening illness" &
          standard_concept_name != "Difficulty coping" &
          standard_concept_name != "Lack of exercise" &
          standard_concept_name != "Gets no exercise")
```

#### Vaccination status

Participants' vaccination status was obtained by querying the All of Us Drug Exposures table (Table A.3) for any SARS-CoV-2 vaccine. Participants were then coded as having a "full series" based on the CDC's definition at the time of data cut-off of either (a) two shots of any mRNA vaccine, or (b) one shot of any other type8. Participants with no record of any SARS-CoV-2 vaccine were coded as "not vaccinated."

##### Table A.3. Vaccination concepts

| Standard Concept Name | Standard Concept Code | Type | N* |
|---|---|---|---|
| SARS-CoV-2 (COVID-19) vaccine, mRNA-BNT162b2 0.1 MG/ML Injectable Suspension | 2468235 | mRNA | 17485 |
| SARS-CoV-2 (COVID-19) vaccine, mRNA-1273 0.2 MG/ML Injectable Suspension | 2470234 | mRNA | 8802 |
| SARS-CoV-2 (COVID-19) vaccine, mRNA spike protein | 2468231 | mRNA | 1084 |
| SARS-COV-2 (COVID-19) vaccine, mRNA, spike protein, LNP, preservative free, 30 mcg/0.3mL dose, tris-sucrose formulation | 217 | mRNA | 985 |
| SARS-COV-2 (COVID-19) vaccine, vector - Ad26 100000000000 UNT/ML Injectable Suspension | 2479835 | protein or vector | 984 |
| SARS-CoV-2 (COVID-19) vaccine, mRNA-1273 0.2 MG/ML | 2470233 | mRNA | 230 |
| SARS-COV-2 (COVID-19) vaccine, vector non-replicating, recombinant spike protein-ChAdOx1, preservative free, 0.5 mL | 210 | protein or vector | 22 |
| SARS-COV-2 (COVID-19) vaccine, vector non-replicating | 2479831 | protein or vector | 18 |

| | | | |
|---|---|---|---|
| SARS-COV-2 (COVID-19) vaccine, UNSPECIFIED | 213 | protein or vector | 17 |
| [Unspecified] | OMOP5048605 | protein or vector | 1 |
| SARS-COV-2 (COVID-19) vaccine, mRNA, spike protein, LNP, preservative free, 10 mcg/0.2mL dose, tris-sucrose formulation | 218 | mRNA | 1 |
| SARS-COV-2 COVID-19 Inactivated Virus Non-US Vaccine Product (BIBP, Sinopharm) | 510 | protein or vector | 1 |
| SARS-CoV-2 (COVID-19) vaccine, mRNA spike protein Injectable Suspension | 2468234 | mRNA | 1 |
| * = Among all program participants | | | |

Table Caption: Standard concept names, codes, vaccine type, and counts of vaccination data queried in *All of* Researcher Workbench dataset builder, Drug Encounters OMOP Table. Raw counts are from the unfiltered (N=104,993) sample of every vaccination encounter.

### Appendix B. Functional Status Indicators

#### Occupational therapy evaluation – Centers for Medicare and Medicaid codes.

The Centers for Medicare and Medicaid Services (CMS) maintains codes and definitions for clinicians to record the medical necessity of services and procedures in the medical record. The codes that record evaluation by an occupational therapist are cataloged under the CPT code directory and are 97165-97168 (see below). These codes only occur in the medical record of a patient with concerns for "performance deficits (i.e., relating to physical, cognitive, or psychosocial skills) that result in activity limitations and/or participation restrictions"9 (emphasis added). This indicator of functional status is therefore highly specific – it is not documented for a person who does not report concern for and/or present with functional performance deficits.
Three codes are available to document the complexity of the clinician's evaluation to determine performance deficits and the therapeutic plan of care. One additional code documents the need for re-evaluation, frequently used when the patient has a decrease in status (due to, e.g., surgical procedure, poor prognosis, and/or complication of medical conditions). CMS defines these as follows:

**97165: Occupational therapy evaluation, low complexity**, requiring these components: An occupational profile and medical and therapy history, which includes a brief history including review of medical and/or therapy records relating to the presenting problem; …, requiring these components: An occupational profile and medical and therapy history, which includes a brief history including review of medical and/or therapy records relating to the presenting problem; An assessment(s) that identifies 1-3 performance deficits (relating to physical, cognitive, or psychosocial skills) that result in activity limitations and/or participation restrictions; and Clinical decision making of low complexity, which includes an analysis of the occupational profile, analysis of data from problem-focused assessment(s), and consideration of a limited number of treatment options. Patient presents with no comorbidities that affect occupational performance. Modification of tasks or assistance (eg, physical or verbal) with assessment(s) is not necessary to enable completion of evaluation component. Typically, 30 minutes are spent face-to-face with the patient and/or family.

**97166 Occupational therapy evaluation, moderate complexity**, typical time with patient 45 minutes, Occupational therapy evaluation, moderate complexity, typical time with patient's family 45 minutes, Occupational therapy evaluation, moderate complexity, requiring these components:
An occupational profile and medical and therapy history, which includes an expanded review of medical and/or therapy records and additional review of physical, cognitive,... , Occupational therapy evaluation, moderate complexity, typical time with patient and family 45 minutes, OCCUPATIONAL THERAPY EVAL MOD COMPLEX 45 MINS, Occupational therapy evaluation, moderate complexity, requiring these components:
An occupational profile and medical and therapy history, which includes an expanded review of medical and/or therapy records and additional review of physical, cognitive, or psychosocial history related to current functional performance; An assessment(s) that identifies 3-5 performance deficits (ie, relating to physical, cognitive, or psychosocial skills) that result in activity limitations and/or participation restrictions; and Clinical decision making of moderate analytic complexity, which includes an analysis of the occupational profile, analysis of data from detailed assessment(s), and consideration of several treatment options. Patient may present with comorbidities that affect occupational performance. Minimal to moderate modification of tasks or assistance (eg, physical or verbal) with assessment(s) is necessary to enable patient to complete evaluation component. Typically, 45 minutes are spent face-to-face with the patient and/or family.

**97167 Occupational therapy evaluation, high complexity**, requiring these components: An occupational profile and medical and therapy history, which includes review of medical and/or therapy records and extensive additional review of physical, cognitive, or psychosocial history related to current functional performance; An assessment(s) that identifies 5 or more performance deficits (ie, relating to physical, cognitive, or psychosocial skills) that result in activity limitations and/or participation restrictions; and Clinical decision making of high analytic complexity, which includes an analysis of the patient profile, analysis of data from comprehensive assessment(s), and consideration of multiple treatment options. Patient presents with comorbidities that affect occupational performance. Significant modification of tasks or assistance (eg, physical or verbal) with assessment(s) is necessary to enable patient to complete evaluation component. Typically, 60 minutes are spent face-to-face w..., Occupational therapy evaluation, high complexity, typical time with patient and family 60 minutes, Occupational therapy evaluation, high complexity, requiring these components: An occupational profile and medical and therapy history, which includes review of medical and/or therapy records and extensive additional review of physical, cognitive, or ps... | [Health Care Activity] - [Therapeutic or Preventive Procedure], Occupational therapy evaluation, high complexity, typical time with patient 60 minutes, Evaluation of occupational therapy established plan of care, typically 60 minutes, TYPICALLY, 60 MINUTES ARE SPENT FACE-TO-FACE WITH THE PATIENT AND/OR FAMILY.

**97168 Re-evaluation of occupational therapy established plan of care,** requiring these components: An assessment of changes in patient functional or medical status with revised plan of care; An update to the initial occupational profile to reflect changes in... | [Health Care Activity] - [Therapeutic or Preventive Procedure], Occupational therapy re-evaluation of established plan of care, typical time with patient 30 minutes, Re-evaluation of occupational therapy established plan of care, typically 30 minutes, Occupational therapy re-evaluation of established plan of care, typical time with patient and family 30 minutes, Re-evaluation of occupational therapy established plan of care, requiring these components: An assessment of changes in patient functional or medical status with revised plan of care; An update to the initial occupational profile to reflect changes in condition or environment that affect future interventions and/or goals; and A revised plan of care. A formal reevaluation is performed when there is a documented change in functional status or a significant change to the plan of care is required. Typically, 30 minutes are spent face-to-face with the patient and/or family.

#### "Finding of functional performance and activity" codes

The Standard medical concept "Finding of Functional Performance and Activity" (OMOP code 4089214, SNOMED code 258636006) was queried for all participants. The findings under this code hierarchy include observations documented by rehabilitation and respiratory therapy providers and based on evaluation findings by the providers. For modelling, these were collapsed into an ordinal variable with three categories roughly corresponding to ranges of functional impairment that may be inferred from these codes. These categories were as follows:
"No functional performance impairment" was given for normal" or "independent" findings, as well as all participants with "NA" values indicating no need for medical care for or observations of functional impairment.
"Some functional performance impairment" was given for codes that may have been documented for a range of functional levels. For instance "Dependence on wheelchair" could be true of a person with acute change in mobility that affects their daily functioning profoundly, and could also be true of a person who had mastered wheelchair mobility and performed their daily routines without assistance. This suggests the important caveat for interpretation that this level has high heterogeneity and low resolution for small increments of difference in level of performance.
"Dependent on others for care" was given for codes which explicitly describe dependence (e.g. "Bed-ridden") and those typically only given in critical care settings (e.g. respiratory dependence codes).

The following dplyr code was used to group these observations:

```
dplyr::mutate(pre_functional_performance_dx = forcats::fct_na_value_to_level
    (pre_functional_performance_dx, "None")) |>
    dplyr::mutate(pre_functional_performance_dx_factor =
    fct_collapse(pre_functional_performance_dx,
         "No Functional Performance Difficulty" = c("None",
            "Get up and go test - normal",
            "Exercises regularly"),
         "Some Functional Performance Difficulty" = c(
    "Difficulty walking",
                                        "Reduced mobility",
                                        "Impaired mobility",
                                        "Physical activity finding",
                                        "Dependence on supplemental oxygen",
                                        "Wheelchair bound",
                                        "Caregiver role strain",
                                        "Dependence on wheelchair",
                                        "Finding related to ability to mobilize",
```

```
                                        "Finding related to ability to move",
                                     "Finding of functional performance and activity",
                                        "Disorders of attention and motor control",
                                        "Walking disability",
                                        "Activity exercise pattern",
                                      "Finding related to ability to cope with pain",
                                        "Difficulty producing voiced sounds",
                                        "Loss of voice",
                                        "Activity intolerance",
                                        "Fine motor impairment"),
     "Dependent on Others for Care" = c(
"Dependence on aspirator",
                                        "Dependence on enabling machine or device",
                                        "Dependence on respirator",
                                        "Bed-ridden",
                                        "Dependence on respiratory device",
                                        "Dependence on ventilator",
                                        "Unable to mobilize"))) |>
dplyr::mutate(pre_functional_performance_dx_factor =
fct_relevel(pre_functional_performance_dx_factor,
                               "No Functional Performance Difficulty",
                               "Some Functional Performance Difficulty",
                               "Dependent on Others for Care"))
```

### Appendix C: Sensitivity analysis for Quantitative bias analysis of cohort discovery

#### 1. Enrollment pre- vs. post-first infection:

Balance between the two groups' distributions of factors which could affect the development of long COVID was assessed using non-parametric propensity score matching in the MatchIt R package.10 The binary outcome of pre-illness versus post-illness enrollment was regressed on COVID variant, primary demographics of race and sex at birth, and the socioeconomic variable of education level. To ensure a match was attempted for all cases, matching without replacement was used. A generalized linear model with logit link functions was fitted with 'nearest' matching. Model fit was assessed with standardized difference in means and plots.

##### *Model*

```
match <- matchit(enrollment_date ~ variant
+ race
+ sex_at_birth
+ Highest_grade_completed,
                data = full_data2, method = "nearest", distance = "glm",
                ratio = 1,
                replace = FALSE)
```

##### *Result*

Pre-infection enrollees were fitted as the control (n=16815) compared to post-infection fitted as cases (n=66971). Most of the sample enrolled prior to their earliest indication of COVID-19. For the unmatched samples, substantial standard mean differences (>0.1) were seen for overall distance (0.32), later omicron variant period (B2-B5) (0.15), Black or African American (0.2331) and White (-0.2056) race, and having completed some or all of high school (0.1174). After balancing, standard mean differences generally increased (grew further from zero), indicating that a good balance on these covariates between pre- and post-illness enrollees could not be achieved.

Overlaps between the pre-infection ('control') and post-infection ('treated') groups is visualized in Figure S.C.1. There is about 45% overlap between the matched post-infection cases and the matched pre-infection controls, and some variables (notably race) could not be matched to achieve. This relatively low match overall suggests that these two groups may be different across one or more important population parameters, which may have been related to programmatic influences on recruitment that affect the sample over time such as adjustments in recruitment to increase sample diversity or the addition of new study sites in previously under-represented regions or patient populations.

#### Figure C.1.

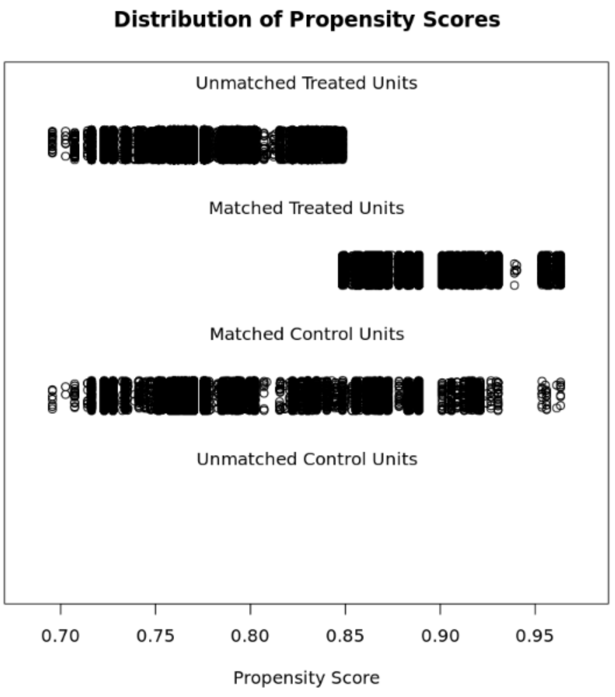

Figure C.1. Caption: Overlaps in propensity for being in the long COVID group between the unmatched versus matched pre-infection enrollees (‘control’) and post-infection enrollees (‘treated’; excluded for the present study’s analysis).

#### Figure C.2

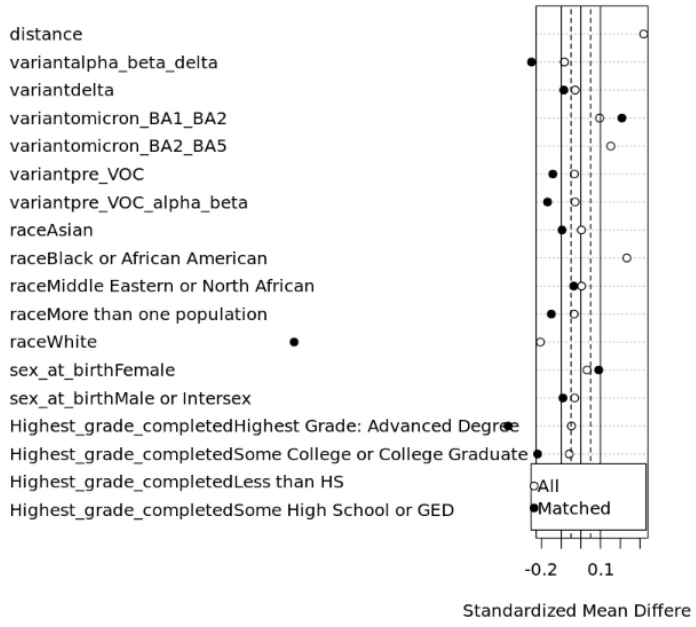

Fig. C.2. Caption: Differences between standardized means of variant, race, sex, and education between participants who joined All of Us before versus after their first infection. Large differences are observed in race and education, with an overall distance of about 0.2 between these groups’ means. These demographic differences most likely reflect period differences in study enrollment efforts targeting under-represented groups in medical research, and may indicate that some of these groups are underrepresented in the present sample.

##### Summary

Sample Sizes:

| | **Control** | Treated |
|---|---|---|
| All | 16815 | 66971 |
| Matched | 16815 | 16815 |
| Unmatched | 0 | 50156 |
| Discarded | 0 | 0 |

Summary of Balance, for All Data compared to Matched Data:

| | All data Std. Mean Difference | Matched data Std. Mean Difference |
|---|---|---|
| Distance | 0.3189 | 1.8529 |
| Variant pre_VOC | -0.0332 | -0.1422 |
| Varian tpre_VOC_alpha_beta | -0.0294 | -0.1691 |
| Variant alpha_beta_delta | -0.0836 | -0.2505 |
| Variant delta | -0.0290 | -0.0863 |
| Variant omicron_BA1_BA2 | 0.0951 | 0.2084 |
| Variant omicron_BA2_BA5 | 0.1522 | 0.8219 |
| Race Asian | 0.0025 | -0.0957 |
| Race Black or African | 0.2331 | 1.6786 |
| Race Middle Eastern or North African | 0.0035 | -0.0361 |
| Race More than one population | -0.0341 | -0.1499 |
| Race White | -0.2056 | -1.4552 |
| sex_at_birth Female | 0.0319 | 0.0908 |
| sex_at_birth Male | -0.0319 | -0.0908 |
| Highest Grade: Advanced Degree | -0.0481 | -0.3692 |
| Some College or College Graduate | -0.0585 | -0.2197 |
| Some High School or GED | 0.1174 | 0.6181 |
| Less than HS | 0.0209 | 0.1560 |

#### 2. Alternative diagnostic duration of symptoms criterion - 90 days versus 28 days since first infection

Using an alternative diagnostic timeframe of 90 days from infection to the earliest mention of long COVID symptoms, n=38,607 participants were classified as long COVID 'cases', and n=3,032 participants who were classified as cases using the diagnostic timeframe of 28 days from infection were re-classified as controls. Adding these to the original n=25,332 participants with no post-infection symptoms gave a total control group of n=28,364. The balance between the long COVID group classification of ≥ long COVID symptom on or after 28 days since first infection was matched on the classification of ≥1 long COVID symptom on or after 90 days since first infection. This was assessed using non-parametric propensity score matching using the MatchIt R package. The binary outcome of having or not having long COVID symptoms at 28-days (the chosen symptom duration for case/control classification in this study) was regressed on the classification at the later 90 day post-infection period, the primary demographics of race and sex at birth, and the socioeconomic variable of education level. To ensure a match was attempted for all cases, matching without replacement was used. A generalized linear model with logit link functions was fitted with 'nearest' matching. Model fit was assessed with standardized difference in means and plots.

##### Model

```
match <- MatchIt::matchit(group ~ lag_class,
                data = join3, method = "nearest", distance = "glm",
                ratio = 1,
                replace = FALSE)
```

##### Result

Overlaps in propensity score between the participants classified as cases at 28 days ('control') and those at 90 days ('treated') is visualized in Figure S.C.3. There was no overlap between cases and controls by the 90-day alternative classification scheme (distance = 1.0).

##### Figure C.3.

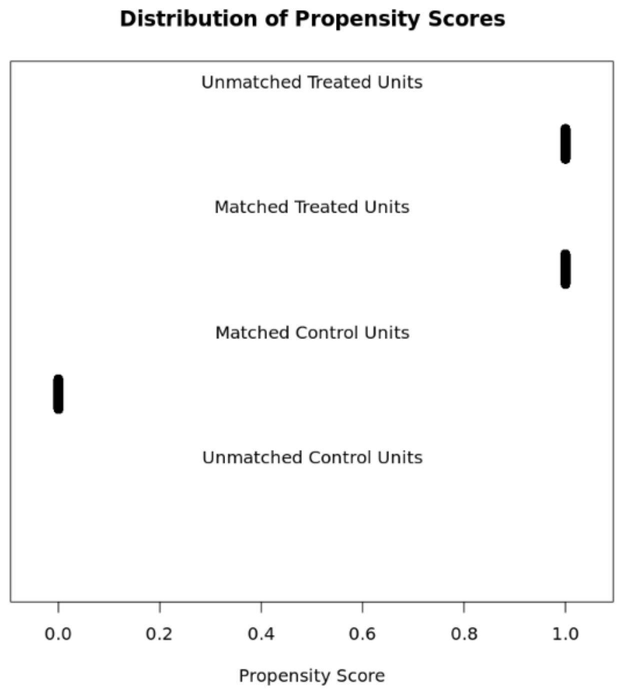

Figure S.C.4 graphs the standard mean differences between the matched samples at the different classification schemes. The close proximity of matched and unmatched participants between the 28-day ("group") and 90 day ("group_90") classification schemes (matched mean differences overlap the unmatched mean differences) suggest that these two classification schemes do not result in substantially different classifications of participants across this sample.

#### Figure C.4

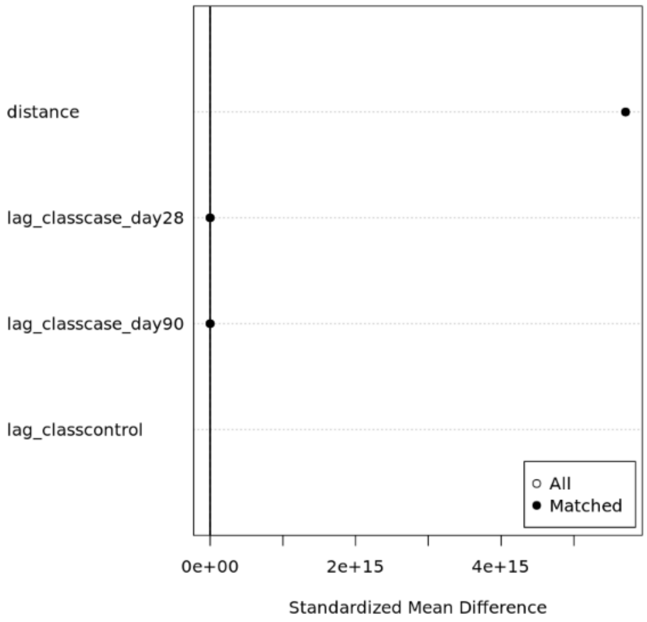

##### *Summary*

Sample Sizes:

| | **Control** | **Treated** |
|---|---|---|
| All | 25332 | 41639 |
| Matched | 25332 | 25332 |
| Unmatched | 0 | 16307 |
| Discarded | 0 | 0 |

Summary of Balance for All Data:

| | Means Treated | Means Control | Std. mean Diff | eCDF Mean | eCDF Max |
|---|---|---|---|---|---|
| Distance | 1.0000 | 0 | 5711494705016479.0 | 0.500 | 1.000 |
| Case at 28 days | 0.0728 | 0 | 0.2802 | 0.073 | 0.073 |
| Case at 90 days | 0.9272 | 0 | 3.5684 | 0.927 | 0.927 |
| Control | 0.0000 | 1.0000 | N/A | 1.000 | 1.000 |

Summary of Balance for Matched Data:

| | Means Treated | Means Control | Std. mean Diff | eCDF Mean | eCDF Max | Standard Pair Distribution |
|---|---|---|---|---|---|---|
| Distance | 1.0000 | 0 | 5711494705016477.0 | 0.5000 | 1.0000 | 5711494705016478.0 |
| Case at 28 days | 0.0687 | 0 | 0.2645 | 0.0687 | 0.0687 | 0.2645 |

| | | | | | | |
|---|---|---|---|---|---|---|
| Case at 90 days | 0.9373 | 0 | 3.5841 | 0.9313 | 0.9313 | 3.5841 |
| Control | 0.0000 | 1 | N/A | 1.0000 | 1.0000 | N/A |

### Appendix D. Model Fitting Notes.

#### Model fitting procedure

Model fitting processes recommended by Harrell 11 and Andersen & Skovgaard 12 were followed for binomial logistic regression with a logit link function for continuous, ordinal, and nominal variables (Table S.D.1). First, a large model was fitted including all variables found in prior literature to be related to long COVID risk or outcomes. This was fitted with natural (restricted) cubic splines for continuous variables, and interaction terms for age with baseline survey responses for performance of physical daily activities and mental/cognitive health, as well as approximate baseline level of functional impairment per EMR entries of "Finding of Functional Activity and Performance" (see Table S.A.1). Modelling and hypothesis testing was completed using the rms package 13. Using $n/15$ or $n/20$ observations per regression coefficient as a guide, we found degrees of freedom in our starting model, indexed to the smaller control group, to be N=24,809, yielding 24,809/20 = 1,240 degrees of freedom.

##### Table D.1. Variables for model

| Type | Covariate | Sources | Model index level/value** |
|---|---|---|---|
| Demographics | Age<br>Sex at birth<br>Race<br>Ethnicity | Self-reported "Basics" survey responses at enrollment<br><br>EMR | Age = 62<br>Female or intersex<br>White<br>Not Hispanic/Latino |
| Acquired demographics/SDH | Education (highest level completed) | Self-reported "Basics" survey responses | Some College |
| SARS-CoV-2 Variant | The calendar period between the starts of major variants of concern in which the first infection occurred. | Self-report of symptoms on the COPE surveys<br><br>EMR condition and measurement codes (SNOMED, OMOP, and ICD-10 vocabularies). | Pre-VOC period |
| Vaccination with full series*** | Whether vaccination with full series (2 shots mRNA, 1 shot other types). | EMR drug encounter codes | Not vaccinated |

| | | | |
|---|---|---|---|
| Pre-infection health and symptoms | Pre-infection total number of long COVID symptom categories with at least one incidence*<br><br>Self-Reported mental health and cognition<br><br>Self-Reported ability to perform physically-demanding daily activities<br><br>Self-Reported social role performance and satisfaction (composite score summing two ordinal items, values 2(~excellent)-10(~very poor) | Self-reported "Overall Health" survey responses at enrollment<br><br>EMR condition codes (SNOMED, OMOP, and ICD-10 vocabularies). | Total number of symptoms with any pre-infection occurrence = 4 (median)<br><br>Individual symptom categories = 0 (no occurrences) |
| Pre-infection daily Functioning | Pre-infection total number of occupational therapy CPT codes*<br><br>Incidences of at least one pre-infection functional performance finding diagnostic codes*, collapsed into three levels. | EMR condition and procedure codes (SNOMED, OMOP, and ICD-10 vocabularies). | CPT total = 0<br><br>"Finding of Functional Performance" impairment level = "None" |
| Note. * = Entered in EHR between five years and four weeks before first infection date.<br>** = Mean, median, or most frequently observed categorical level.<br>***Four different fittings of vaccination data were tested in the final model. These were (a) three-level factor with *Before first infection* (n=886), *After first infection* (n=2,449), and *Not vaccinated* by data cut-off (July 2022) (n=63635), (b) datetime (month_year) of full series vaccination, and (c) lag time between first infection and full series vaccination. Due to the small number of people with vaccination at the time of this dataset's cut-off, these models were overparameterized against the primary covariate of functional status and exhibited larger error per AIC delta. These were thus judged to be overfitted. The best fit to these data for the present study's aims was found for the binary fitting of this predictor. | | | |

#### Summary of final fit

The optimal model balance between $R^2$ and BIC delta (smaller value) was found for an additive model with linear fits for age and pre-infection CPT unit count and a restricted cubic spline with four knots for pre-infection prevalence of long COVID symptoms. For this paper's aim, parsimoniousness was favored over accuracy for a simple descriptive model.

The final model's moderate $R^2$ indicates that there is more variance in the outcome than can be explained by the model. This is unsurprising given the evolving nature of both the disease and humans' identification of it during the era in question, and the non-specific dataset used (a general national biobank, not a purpose-built COVID19 study cohort).

Several aspects of this model may have contributed to unexplained variance. First, model fitting decisions served associative (as opposed to predictive) aim, which provide that a high $R^2$ is not required to believe the relationships observed. We therefore assessed model fit using the AIC, rather than the more conservative BIC and maximized $R^2$ as we would for a predictive model. Second, some parameters had high variance inflation factors (see Table D.4), primarily due to the inclusion of individual variables and summary variables of which the individual variables were components (e.g. symptom total and individual symptoms tallied under that total). Excessive variance due to bias is not suspected since the residual plots, which show whether there is bias in the distribution of residuals, are fairly regular.

##### Table D.2. Final Model Summary

| | Model Likelihood Ratio Test | Discrimination Indexes | Rank Discrim. Indexes |
|---|---|---|---|
| Obs 65464 | LR chi2 26194.58 | $R^2$ 0.449 | C 0.850 |
| FALSE 24809 | d.f. 78 | $R^2$ (78,65464)0.329 | Dxy 0.699 |
| TRUE 40655 | Pr(> chi2) <0.0001 | $R^2$ (78,46221.3)0.432 | gamma 0.699 |
| max \|deriv\| 3e-08 | | Brier 0.150 | tau-a 0.329 |

Table D.2. Caption: The final model had 78 degrees of freedom from the fitted parameters. Model discrimination was good, with a maximum likelihood ratio $X^2$ of 26194.58 and an area under the receiver operating curve (C) of 85%. The $R^2$ – total variance explained by the fitted model – was 45%, indicating about half of the variance was unexplained by this model. Given the large sample size and the highly significant $X^2$ of the maximum likelihood ratio ($p$ <0.0001), this model was judged to have adequate fit for determining association.

##### Table D.3. ANOVA omnibus test of effects

| Covariate | Chi-Square | d.f. | P |
|---|---|---|---|
| variant | 5832.59 | 5.00 | 0.00 |
| vaccination2 | 60.13 | 1.00 | 0.00 |
| age (Factor+Higher Order Factors) | 148.52 | 12.00 | 0.00 |
| All Interactions | 52.03 | 11.00 | 0.00 |
| sex_at_birth | 11.67 | 1.00 | 0.00 |
| race | 19.41 | 4.00 | 0.00 |
| ethnicity | 4.43 | 1.00 | 0.04 |
| Highest_grade_completed | 10.52 | 5.00 | 0.06 |
| pre_infection_sx_total (Factor+Higher Order Factors) | 1636.72 | 8.00 | 0.00 |
| All Interactions | 44.63 | 4.00 | 0.00 |
| Nonlinear (Factor+Higher Order Factors) | 1630.38 | 6.00 | 0.00 |
| social_scale | 13.39 | 8.00 | 0.10 |
| Rate_your_mental_health | 18.85 | 5.00 | 0.00 |
| Can_you_complete_daily_activities (Factor+Higher Order Factors) | 17.65 | 10.00 | 0.06 |
| All Interactions | 8.40 | 5.00 | 0.14 |
| pre_functional_performance_dx_factor (Factor+Higher Order Factors) | 22.15 | 4.00 | 0.00 |
| All Interactions | 0.91 | 2.00 | 0.63 |
| pre_abdominal_dx | 0.13 | 1.00 | 0.72 |
| pre_anxiety_dx | 2.86 | 1.00 | 0.09 |
| pre_chestpain_dx | 0.03 | 1.00 | 0.86 |

| | | | |
|---|---|---|---|
| pre_cough_dx | 0.22 | 1.00 | 0.64 |
| pre_depression_dx | 2.15 | 1.00 | 0.14 |
| pre_diarrhea_dx | 0.15 | 1.00 | 0.70 |
| pre_dizziness_dx | 0.05 | 1.00 | 0.83 |
| pre_dyspnea_dx | 0.50 | 1.00 | 0.48 |
| pre_fatigue_dx | 0.43 | 1.00 | 0.51 |
| pre_fever_dx | 0.36 | 1.00 | 0.55 |
| pre_headache_dx | 0.00 | 1.00 | 0.97 |
| pre_cognition_dx | 0.01 | 1.00 | 0.94 |
| pre_joint_pain_dx | 2.32 | 1.00 | 0.13 |
| pre_me_cfs_dx | 0.05 | 1.00 | 0.83 |
| pre_menstruation_dx | 0.15 | 1.00 | 0.70 |
| pre_muscle_pain_dx | 0.66 | 1.00 | 0.42 |
| pre_msk_chest_pain_dx | 0.21 | 1.00 | 0.64 |
| pre_palpitations_dx | 0.57 | 1.00 | 0.45 |
| pre_paraesthesia_dx | 0.22 | 1.00 | 0.64 |
| pre_sexual_function_dx | 0.00 | 1.00 | 0.99 |
| pre_rash_dx | 1.71 | 1.00 | 0.19 |
| pre_sleep_dx | 5.47 | 1.00 | 0.02 |
| pre_tachycardia_dx | 0.71 | 1.00 | 0.40 |
| age * Can_you_complete_daily_activities (Factor+Higher Order Factors) | 8.40 | 5.00 | 0.14 |
| age * pre_functional_performance_dx_factor (Factor+Higher Order Factors) | 0.91 | 2.00 | 0.63 |
| age * pre_infection_sx_total (Factor+Higher Order Factors) | 44.63 | 4.00 | 0.00 |
| Nonlinear | 26.12 | 3.00 | 0.00 |
| Nonlinear Interaction : f(A,B) vs. AB | 26.12 | 3.00 | 0.00 |
| TOTAL NONLINEAR | 1630.38 | 6.00 | 0.00 |
| TOTAL INTERACTION | 52.03 | 11.00 | 0.00 |
| TOTAL NONLINEAR + INTERACTION | 1643.31 | 14.00 | 0.00 |
| TOTAL | 15819.26 | 76.00 | 0.00 |

#### Figure D.1. Contributions of each variable to model effects.

Plot showing the contribution of each variable ($X^2$) to the model's omnibus effect size. Covariates are plotted on the *Y* axis; the least-significant contributor is at the top, and the rest are plotted down the axis in order of effect to the most-significant contributor at the bottom. The effect size is plotted on the *X* axis as $X^2$ minus the degrees of freedom. Each variable's effect is plotted as a dot intersecting the covariate with its effect size value. The top (smallest) contributor is the interaction between age and prior functional performance ($X^2$=0.0, $p$=0.994); the bottom (largest) contributor is infection variant, ($X^2$=5832.6, $p$<0.0000).

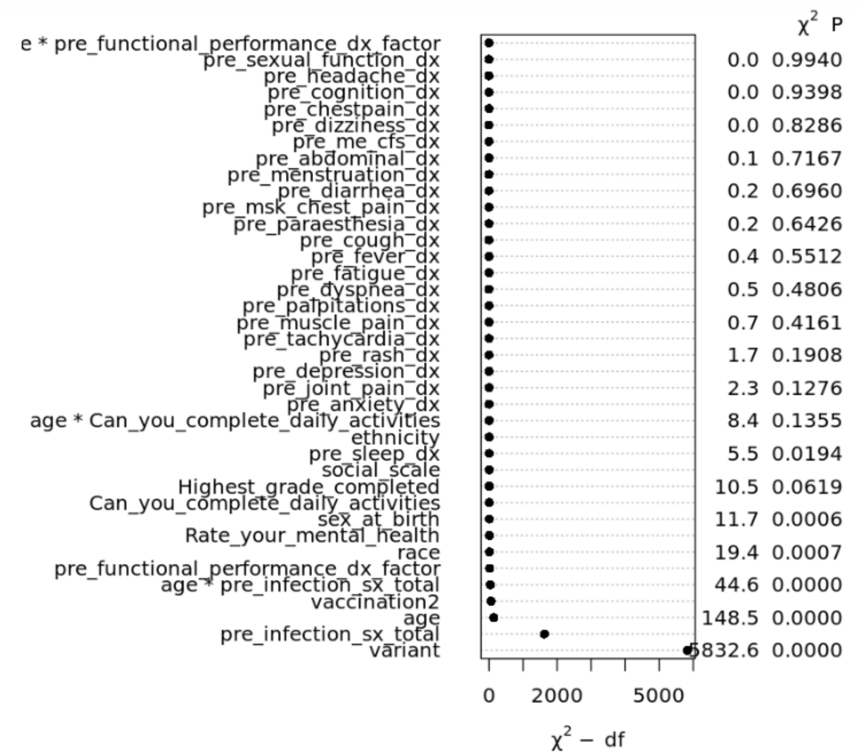

Figure D.1. Caption: Plot showing the contribution of each variable ($X^2$) to the model's omnibus effect size. Covariates are plotted on the *Y* axis; the least-significant contributor is at the top, and the rest are plotted down the axis in order of effect to the most-significant contributor at the bottom. The effect size is plotted on the *X* axis as $X^2$ minus the degrees of freedom. Each variable's effect is plotted as a dot intersecting the covariate with its effect size value. The top (smallest) contributor is the interaction between age and prior functional performance ($X^2$=0.0, $p$=0.994); the bottom (largest) contributor is infection variant, ($X^2$=5832.6, $p$<0.0000).

#### Summary of fitting procedure, structured tests, and findings

##### *Initial full model*

```
initial_full <- rms::lrm(group == "Long COVID" ~
                variant +
                vaccination2 +
                 rcs(age,7) +
                 sex_at_birth +
                 race +
                 ethnicity +
                 Highest_grade_completed +
                 rcs(pre_infection_sx_total,4) +
                 rcs(cpt_total_precovid,5) +
                 pre_functional_performance_dx_factor +
                 social_scale +
                 Rate_your_mental_health +
                 Can_you_complete_daily_activities +
                      pre_abdominal_dx +
                      pre_anxiety_dx +
                      pre_chestpain_dx +
```

```
            pre_cough_dx +
            pre_depression_dx +
            pre_diarrhea_dx +
            pre_dizziness_dx +
            pre_dyspnea_dx +
            pre_fatigue_dx +
            pre_fever_dx +
            pre_headache_dx +
            pre_cognition_dx +
            pre_joint_pain_dx +
            pre_me_cfs_dx +
            pre_menstruation_dx +
            pre_muscle_pain_dx +
            pre_msk_chest_pain_dx +
            pre_palpitations_dx +
            pre_paraesthesia_dx +
            pre_sexual_function_dx +
            pre_rash_dx +
            pre_sleep_dx +
            pre_tachycardia_dx +
          rcs(age,7):Can_you_complete_daily_activities +
          rcs(age,7):Rate_your_mental_health +
          rcs(age,7):pre_functional_performance_dx_factor,
      data = temp3, y=TRUE, x=TRUE)
```

#### *Structured model tests of complexity*

##### Continuous variable splines and linearity tests

We tested the initial full model against simplified models with reduced spline knots for age, pre-infection symptoms, and number of pre-infection CPT billing units for occupational therapy. There was very strong evidence, provided by smaller observed BIC values compared to initial and all other linearity permutations, to support a model with a linear fits for age and pre-infection CPT billing units, and a restricted cubic spline with five knots for pre-infection number of symptoms.

##### Checking interaction terms: comparing to additive models

All interaction terms in the working initial model were dropped, then added back into the model singly. There was strong evidence from the BIC delta values (compared to the working model and all other combinations of working model interaction terms) for a model with interactions between age and the pre-infection variables of (a) physical ability to complete daily activities, (b) EHR-derived functional performance level, and (c) number of incident symptoms. The largest BIC delta from all iterations of these comparisons was the model with the age:self-reported MH dropped. All other interaction terms were retained.

##### Test for main effects

Alternative models were fitted with main effects for pre-infection total number of symptoms, pre-infection functional performance factor, variant of concern at time of first infection, and vaccination status at time of first infection. There was no evidence of significant differences between these models and the working model (BIC delta < |5|). The working model was retained for fit checking.

##### Model fit checks

There were no significant differences observed in residual patterns between the long COVID and Recovered groups. Residual fits on continuous variables are shown in Fig.s S.D.2-S.D.4. Visual evidence of model fit was satisfactory for these data.

Figure D.2. Residual fit of age

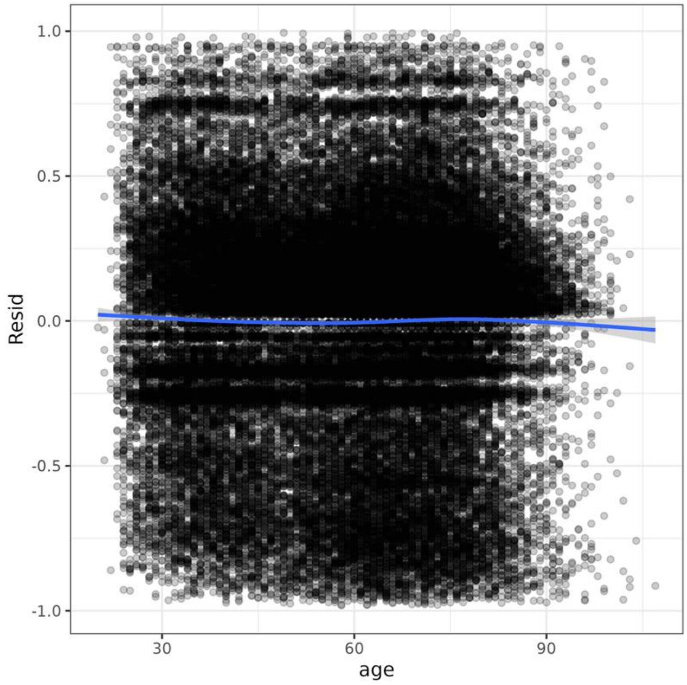

Figure D.3. Residual fits of total number of occupational therapy CPT units billed in five years preceding infection

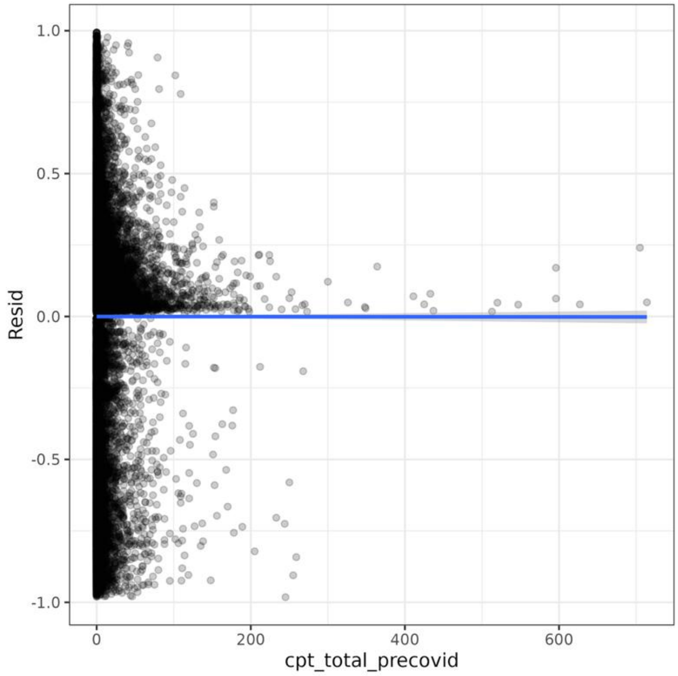

Figure D.4. Residual fits of total number of long COVID symptom/condition types occurring prior to infection

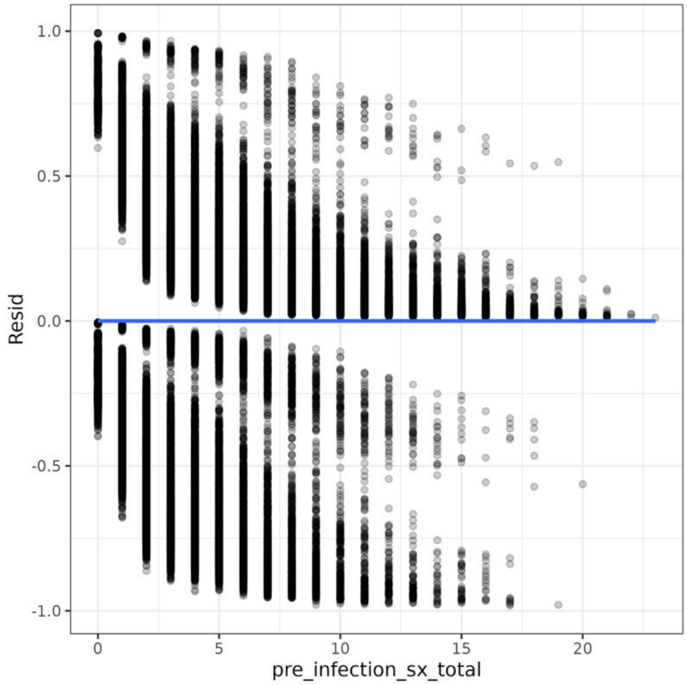

*Final fitted model:*

```
final_fit <- rms::lrm(group == "Long COVID" ~
                variant +
                vaccination2 +
                age +
                sex_at_birth +
                race +
                ethnicity +
                Highest_grade_completed +
                rcs(pre_infection_sx_total,5) +
                social_scale +
                Rate_your_mental_health +
                Can_you_complete_daily_activities +
                pre_functional_performance_dx_factor +
                cpt_total_precovid +
                      pre_abdominal_dx +
                      pre_anxiety_dx +
                      pre_chestpain_dx +
                      pre_cough_dx +
                      pre_depression_dx +
                      pre_diarrhea_dx +
                      pre_dizziness_dx +
                      pre_dyspnea_dx +
                      pre_fatigue_dx +
                      pre_fever_dx +
                      pre_headache_dx +
                      pre_cognition_dx +
```

```
            pre_joint_pain_dx +
            pre_me_cfs_dx +
            pre_menstruation_dx +
            pre_muscle_pain_dx +
            pre_msk_chest_pain_dx +
            pre_palpitations_dx +
            pre_paraesthesia_dx +
            pre_sexual_function_dx +
            pre_rash_dx +
            pre_sleep_dx +
            pre_tachycardia_dx +
          age:Can_you_complete_daily_activities +
          age:pre_functional_performance_dx_factor +
          age:rcs(pre_infection_sx_total,5),
      data = temp3, y=TRUE, x=TRUE)
```

##### *Assessment of collinearity of model covariates*

The collinearity of all covariates in the model are reported below.

##### Variance inflation factor (VIF) of fitted covariates

The variance inflation factor (VIF) scores for the fitted model covariates was assessed with the rms::vif function[13] in R. This estimates the percentage of extra variance in each variable's impact upon the development of at least one long COVID symptom as a function of the model $R^2$ that may be expected for each variable if they are strongly correlated with the outcome or other variables in the model. VIF approximates the precision of each variable's slope; values at or near 1 indicate no concern for collinearity.[14,15]

Based on the exploratory aims of this study, the binary outcome variable, the large sample size, and the assumption that certain health conditions in long COVID frequently co-occur, the a-priori threshold for variance inflation we chose was a liberal 10.0. This was not exceeded by any variables in this model, but was nearly met by pre-infection incidence of ME/CFS (9.52), joint pain (8.05), and the interaction between number of pre-infection symptoms and age (8.84 – 7.29). There were also a number variables and variable levels between 5 and 10. (Table D.4).

Multicollinearity tends to reduce the estimate of a variable's true significance and increases its confidence interval, thus prior ME/CFS, joint pain, and the interaction between age and prior number of symptoms may have been underestimated as risk factors for long COVID in this model. This model's collinearity may be explained by several factors. First is the difficulty classifying long COVID symptom incidences as meaningfully different from pre-infection symptom experiences or presentations. For instance, pre-infection ME/CFS was relatively rare in this sample, however as it was one of the post-infection symptoms we used to classify as cases it is possible that people with ongoing ME/CFS (but without exacerbation or change in their condition after infection) were inappropriately classified as "cases". Misclassification may have also accounted for joint pain's higher collinearity. Considering its high prevalence in the entire sample during the five-year look-back from date of first infection, this may have been a less discriminating symptom. The complexity of the relationships between health changes and aging is not unexpected, but its impact on risk is critical to understand and warrants a deeper examination in a future study.

##### Table D.4. Variance inflation factors of all covariates and interaction terms

| **Covariate** | **Variance inflation factor** |
|---|---|
| variant=Pre-VOC, Alpha, Beta | 1.238834e+00 |
| variant=Alpha, Beta, Delta | 1.132717e+00 |

| | |
|---|---|
| variant=Delta | 1.176021e+00 |
| variant=Omicron BA1-BA2 | 1.200145e+00 |
| variant=Omicron BA2-BA5 | 1.113494e+00 |
| vaccination=Full Series | 1.021039e+00 |
| Age | 6.049463e+00 |
| sex_at_birth=Male | 1.128947e+00 |
| race=Black or African American | 5.895579e+00 |
| race=Middle Eastern or North African | 1.212751e+00 |
| race=More than one population | 1.583254e+00 |
| race=White | 6.164593e+00 |
| ethnicity=Not Hispanic or Latino | 1.019674e+00 |
| Highest_grade_completed=College or college grad | 1.494630e+00 |
| Highest_grade_completed=High School/GED | 1.570400e+00 |
| Highest_grade_completed=Some High School | 1.229526e+00 |
| Highest_grade_completed=Less than high school | 1.044473e+00 |
| Highest_grade_completed=No Answer | 1.060842e+00 |
| pre_infection_sx_total | 4.144329e+03 |
| pre_infection_sx_total' | 6.960308e+04 |
| pre_infection_sx_total'' | 1.413850e+05 |
| pre_infection_sx_total''' | 1.974925e+04 |
| social_scale=3 | 1.831219e+00 |
| social_scale=4 | 2.475786e+00 |
| social_scale=5 | 2.278675e+00 |
| social_scale=6 | 2.530618e+00 |
| social_scale=7 | 1.994741e+00 |
| social_scale=8 | 1.915202e+00 |
| social_scale=9 | 1.449554e+00 |
| social_scale=10 | 1.351644e+00 |
| Rate_your_mental_health=Very Good | 2.083809e+00 |
| Rate_your_mental_health=Good | 2.512260e+00 |
| Rate_your_mental_health=Fair | 2.290396e+00 |
| Rate_your_mental_health=Poor | 1.508990e+00 |
| Rate_your_mental_health=No Answer | 1.051157e+00 |
| Can_you_complete_daily_activities=Mostly | 1.490209e+01 |
| Can_you_complete_daily_activities=Moderately | 1.720550e+01 |
| Can_you_complete_daily_activities=A little | 2.072651e+01 |
| Can_you_complete_daily_activities=Not at all | 1.985066e+01 |
| Can_you_complete_daily_activities=No Answer | 1.777763e+01 |
| pre_functional_performance_dx_factor=Some | 3.912027e+01 |
| pre_functional_performance_dx_factor=Dependent | 3.544598e+01 |
| cpt_total_precovid | 1.052716e+00 |
| pre_abdominal_dx | 6.944247e+01 |
| pre_anxiety_dx | 6.221869e+01 |
| pre_chestpain_dx | 6.856771e+01 |
| pre_cough_dx | 5.960201e+01 |
| pre_depression_dx | 5.786623e+01 |
| pre_diarrhea_dx | 3.670505e+01 |
| pre_dizziness_dx | 4.089403e+01 |
| pre_dyspnea_dx | 5.324548e+01 |
| pre_fatigue_dx | 5.307157e+01 |
| pre_fever_dx | 2.560118e+01 |
| pre_headache_dx | 4.415449e+01 |
| pre_cognition_dx | 2.132864e+01 |

| | |
|---|---|
| pre_joint_pain_dx | 8.048326e+01 |
| pre_me_cfs_dx | 9.517710e+00 |
| pre_menstruation_dx | 2.559214e+01 |
| pre_muscle_pain_dx | 2.436233e+01 |
| pre_msk_chest_pain_dx | 1.347388e+01 |
| pre_palpitations_dx | 2.703584e+01 |
| pre_paraesthesia_dx | 2.203656e+01 |
| pre_sexual_function_dx | 5.349296e+00 |
| pre_rash_dx | 4.722936e+01 |
| pre_sleep_dx | 6.044857e+01 |
| pre_tachycardia_dx | 2.275191e+01 |
| age * Can_you_complete_daily_activities=Mostly | 1.525799e+01 |
| age * Can_you_complete_daily_activities=Moderately | 1.730822e+01 |
| age * Can_you_complete_daily_activities=A little | 2.043648e+01 |
| age * Can_you_complete_daily_activities=Not at all | 1.979777e+01 |
| age * Can_you_complete_daily_activities=No Answer | 1.778441e+01 |
| age * pre_functional_performance_dx_factor=Some | 2.192163e+01 |
| age * pre_functional_performance_dx_factor=Dependent | 2.715921e+01 |
| age * pre_infection_sx_total | 8.839911e+02 |
| age * pre_infection_sx_total' | 7.293169e+04 |
| age * pre_infection_sx_total'' | 1.456008e+05 |
| age * pre_infection_sx_total''' | 1.999367e+04 |

Table D.4. Caption: Variance inflation factor (VIF) for the parameters of each covariate in the model, resulting from several covariates' effects on the outcome increasing or decreasing at the same time and magnitude (such that the effect of one could be predicted by or accounted for by the other). This is an estimate of multicollinearity in the final model. VIF was below 5.0 for most covariates, indicating low risk for multicollinearity. Seventeen parameter values (e.g. levels in a categorical variable or interaction) were above 5. None exceeded 10.

#### Appendix E. Expanded results.

##### Figure E.1. Time of first infection, by long COVID group

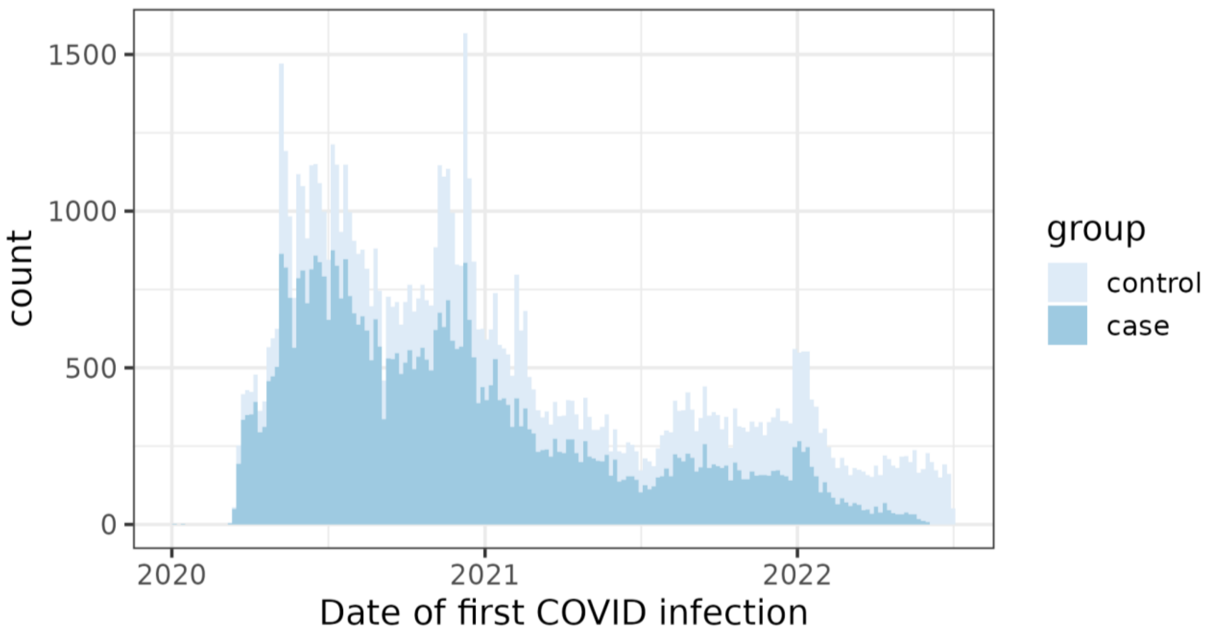

Fig. E.1. Caption: Histogram of the number of participants with (darker blue) versus without (lighter blue) long COVID by first infection date through July 2022. The Y axis plots the number of participants ascending from zero (bottom) to over 1,500 (top); the X axis is a timeline from (left to right) January 1 2020 through July 31 2022. Both groups show a similar profile; they begin with a steep spike in infections about March 2020, a large and prolonged peak between April and September 2020, and briefer peaks between about October 2020 – January 2021 and in January 2022.

#### Figure E.2. Among vaccinated participants, distributions of first infection and first vaccination date.

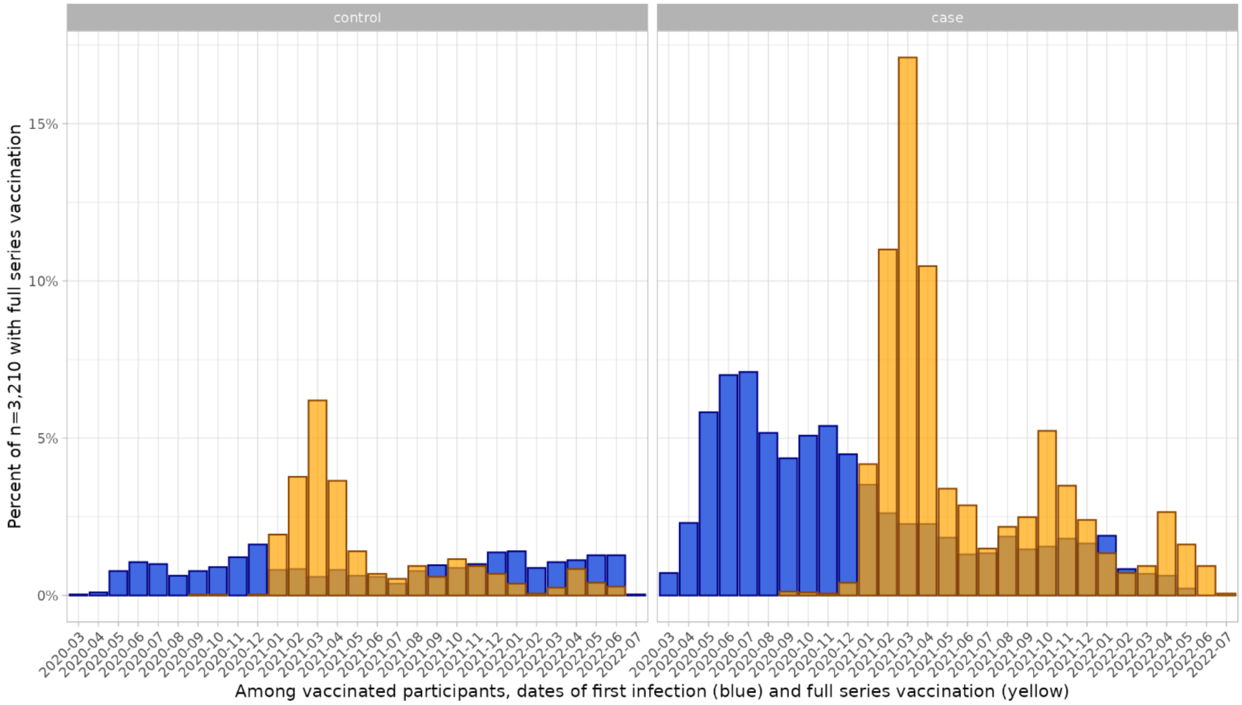

Fig. E.2. Caption: Two side-by-side bar graphs (controls on the left, cases on the right) of the timing of first infection and first full-series vaccination among the n=3,210 participants who were fully vaccinated. This graph suggests that, among participants with full vaccination, those with at least one long COVID symptom at any point had a higher disease incidence through early 2022 (spiking sharply in early 2020 and tapering gradually through 2021), and a higher uptake of both initial vaccination (December 2020 through about June 2021) and boosters (spiking in October 2021 and April 2022).

### Interaction terms of age with pre-infection symptom totals, clinical functional status observation codes, and self-reported ability to complete daily physical activities.

#### Table E.1. Interaction: Age by functional status inferred from pre-infection EHR clinical observations

| Age (years) | Some prior difficulty | Severe prior difficulty |
|---|---|---|
| 25 | 1.06 (0.68, 1.6), 0.80 | 1.21 (0.70, 2.1), <0.49 |
| 45 | 1.03 (0.70, 1.5), 0.87 | 1.31 (0.86, 2.0), <0.21 |
| 65 | 1.01 (0.70, 1.4), 0.98 | 1.42 (0.98, 2.1), 0.07 |
| 75 | 0.98 (0.67, 1.40), 0.92 | 1.54 (1.01, 2.4), <0.04 |

Table E.1. Legend. Adjusted odds by age and pre-infection functional performance level (EHR code under "Finding of Functional Performance and Activity") of developing long COVID) compared to no prior difficulty.

##### Table E.2. Interaction: Age by pre-infection self-reported physical ability.

| Age (years) | Mostly | A little bit | Not at all |
|---|---|---|---|
| 25 | 1.06 (0.93, 1.21), 0.36 | 1.05 (0.84, 1.30), 0.68 | 1.09 (0.66, 1.78), 0.74 |
| 45 | 1.05 (0.98, 1.13), 0.18 | 0.99 (0.87, 1.12), 0.86 | 0.94 (0.71, 1.23), 0.65 |
| 65 | 1.04 (0.98, 1.11), 0.17 | 0.94 (0.85, 1.03), 0.17 | 0.81 (0.66, 0.99), 0.04 |
| 75 | 1.03 (0.94, 1.14), 0.52 | 0.88 (0.75, 1.05), 0.6 | 0.70 (0.48, 1.01), 0.06 |

Table E.2. Legend: Adjusted odds by age and self-rated ability to complete physical daily activities of developing long COVID, compared to “completely” able.

##### Table E.3. Interaction: Age by pre-infection total incident symptoms.

| Age (years) | 0 symptoms | 1 symptom | 7 symptoms |
|---|---|---|---|
| 25 | 0.22 (0.05, 0.90), 0.04 | 0.47 (0.16, 1.37), 0.17 | 1.37 (0.47, 4.02), 0.56 |
| 45 | 0.18 (0.04, 0.74), 0.02 | 0.42 (0.15, 1.23), 0.11 | 1.48 (0.51, 4.31), 0.47 |
| 65 | 0.15 (0.04, 0.61), 0.01 | 0.38 (0.13, 1.11), 0.08 | 1.6 (0.55, 4.65), 0.39 |
| 85 | 0.12 (0.03, 0.50), <0.00 | 0.35 (0.12, 1.01), 0.05 | 1.78 (0.59, 5.04), 0.31 |

Table E.3. Legend. Adjusted odds by age and pre-infection symptom incidences of developing long COVID, compared to the median of four pre-infection symptoms.

#### Pearson's R correlation coefficients between pre-infection symptoms

The correlation matrix used to compute first principal components (Main document, Fig 3) between symptoms is presented here. These values range from zero (menstrual disorders with dyspnea, $R^2$=0.00) to moderate (depression with anxiety, $R^2$=0.53). Most correlations fall below $R^2$=0.31, suggesting a low risk for symptom-wise collinearity.

##### Table E.4. Correlation matrix of pre-infection symptoms

| | abdominal | anxiety | Chest pain | cough | depression | diarrhea | dizziness | dyspnea | fatigue | fever | headache | cognition | joint_pain | me_cfs | menstruation | muscle_pain | msk_chest_pain | palpitations | paraesthesia | sexual_function | rash | sleep | tachycardia |
|---|---|---|---|---|---|---|---|---|---|---|---|---|---|---|---|---|---|---|---|---|---|---|---|
| abdominal | 1.00 | 0.25 | 0.32 | 0.23 | 0.25 | 0.32 | 0.22 | 0.25 | 0.23 | 0.19 | 0.27 | 0.13 | 0.24 | 0.09 | 0.16 | 0.19 | 0.14 | 0.15 | 0.14 | 0.07 | 0.18 | 0.20 | 0.17 |
| anxiety | 0.25 | 1.00 | 0.24 | 0.18 | 0.53 | 0.20 | 0.19 | 0.19 | 0.22 | 0.12 | 0.24 | 0.22 | 0.21 | 0.09 | 0.13 | 0.16 | 0.12 | 0.14 | 0.12 | 0.07 | 0.17 | 0.29 | 0.16 |
| Chest pain | 0.32 | 0.24 | 1.00 | 0.29 | 0.23 | 0.22 | 0.27 | 0.40 | 0.24 | 0.18 | 0.28 | 0.16 | 0.28 | 0.09 | 0.05 | 0.20 | 0.33 | 0.22 | 0.16 | 0.04 | 0.16 | 0.25 | 0.21 |
| cough | 0.23 | 0.18 | 0.29 | 1.00 | 0.19 | 0.20 | 0.20 | 0.33 | 0.22 | 0.24 | 0.22 | 0.12 | 0.26 | 0.08 | 0.04 | 0.17 | 0.11 | 0.13 | 0.12 | 0.03 | 0.18 | 0.21 | 0.16 |
| depression | 0.25 | 0.53 | 0.23 | 0.19 | 1.00 | 0.20 | 0.19 | 0.20 | 0.23 | 0.12 | 0.24 | 0.26 | 0.23 | 0.10 | 0.11 | 0.16 | 0.11 | 0.10 | 0.12 | 0.06 | 0.16 | 0.31 | 0.15 |
| diarrhea | 0.32 | 0.20 | 0.22 | 0.20 | 0.20 | 1.00 | 0.20 | 0.21 | 0.21 | 0.20 | 0.21 | 0.12 | 0.18 | 0.08 | 0.06 | 0.15 | 0.11 | 0.11 | 0.11 | 0.04 | 0.16 | 0.18 | 0.17 |
| dizziness | 0.22 | 0.19 | 0.27 | 0.20 | 0.19 | 0.20 | 1.00 | 0.25 | 0.26 | 0.14 | 0.27 | 0.15 | 0.21 | 0.10 | 0.04 | 0.16 | 0.10 | 0.21 | 0.15 | 0.04 | 0.14 | 0.20 | 0.16 |
| dyspnea | 0.25 | 0.19 | 0.40 | 0.33 | 0.20 | 0.21 | 0.25 | 1.00 | 0.26 | 0.20 | 0.23 | 0.14 | 0.22 | 0.09 | 0.00 | 0.17 | 0.12 | 0.21 | 0.13 | 0.01 | 0.14 | 0.27 | 0.24 |
| fatigue | 0.23 | 0.22 | 0.24 | 0.22 | 0.23 | 0.21 | 0.26 | 0.26 | 1.00 | 0.17 | 0.22 | 0.16 | 0.24 | 0.34 | 0.07 | 0.21 | 0.12 | 0.17 | 0.17 | 0.05 | 0.18 | 0.26 | 0.14 |
| fever | 0.19 | 0.12 | 0.18 | 0.24 | 0.12 | 0.20 | 0.14 | 0.20 | 0.17 | 1.00 | 0.17 | 0.09 | 0.12 | 0.05 | 0.04 | 0.12 | 0.08 | 0.09 | 0.07 | 0.02 | 0.12 | 0.12 | 0.20 |
| headache | 0.27 | 0.24 | 0.28 | 0.22 | 0.24 | 0.21 | 0.27 | 0.23 | 0.22 | 0.17 | 1.00 | 0.17 | 0.21 | 0.08 | 0.12 | 0.21 | 0.13 | 0.15 | 0.16 | 0.06 | 0.16 | 0.20 | 0.17 |
| cognition | 0.13 | 0.22 | 0.16 | 0.12 | 0.26 | 0.12 | 0.15 | 0.14 | 0.16 | 0.09 | 0.17 | 1.00 | 0.13 | 0.07 | 0.01 | 0.10 | 0.06 | 0.06 | 0.09 | 0.02 | 0.09 | 0.17 | 0.11 |
| joint_pain | 0.24 | 0.21 | 0.28 | 0.26 | 0.23 | 0.18 | 0.21 | 0.22 | 0.24 | 0.12 | 0.21 | 0.13 | 1.00 | 0.10 | 0.04 | 0.23 | 0.15 | 0.14 | 0.17 | 0.05 | 0.19 | 0.28 | 0.11 |
| ME/CFS | 0.09 | 0.09 | 0.09 | 0.08 | 0.10 | 0.08 | 0.10 | 0.09 | 0.34 | 0.05 | 0.08 | 0.07 | 0.10 | 1.00 | 0.05 | 0.11 | 0.06 | 0.07 | 0.09 | 0.03 | 0.08 | 0.13 | 0.04 |
| menstruation | 0.16 | 0.13 | 0.05 | 0.04 | 0.11 | 0.06 | 0.04 | 0.00 | 0.07 | 0.04 | 0.12 | 0.01 | 0.04 | 0.05 | 1.00 | 0.05 | 0.04 | 0.05 | 0.05 | 0.10 | 0.11 | 0.02 | 0.06 |
| muscle_pain | 0.19 | 0.16 | 0.20 | 0.17 | 0.16 | 0.15 | 0.16 | 0.17 | 0.21 | 0.12 | 0.21 | 0.10 | 0.23 | 0.11 | 0.05 | 1.00 | 0.16 | 0.11 | 0.16 | 0.06 | 0.14 | 0.18 | 0.10 |
| msk_chest_pain | 0.14 | 0.12 | 0.33 | 0.11 | 0.11 | 0.11 | 0.10 | 0.12 | 0.12 | 0.08 | 0.13 | 0.06 | 0.15 | 0.06 | 0.04 | 0.16 | 1.00 | 0.08 | 0.10 | 0.03 | 0.10 | 0.12 | 0.08 |
| palpitations | 0.15 | 0.14 | 0.22 | 0.13 | 0.10 | 0.11 | 0.21 | 0.21 | 0.17 | 0.09 | 0.15 | 0.06 | 0.14 | 0.07 | 0.05 | 0.11 | 0.08 | 1.00 | 0.10 | 0.03 | 0.10 | 0.13 | 0.18 |
| paraesthesia | 0.14 | 0.12 | 0.16 | 0.12 | 0.12 | 0.11 | 0.15 | 0.13 | 0.17 | 0.07 | 0.16 | 0.09 | 0.17 | 0.09 | 0.05 | 0.16 | 0.10 | 0.10 | 1.00 | 0.03 | 0.11 | 0.14 | 0.07 |
| sexual_function | 0.07 | 0.07 | 0.04 | 0.03 | 0.06 | 0.04 | 0.04 | 0.01 | 0.05 | 0.02 | 0.06 | 0.02 | 0.05 | 0.03 | 0.10 | 0.06 | 0.03 | 0.03 | 0.03 | 1.00 | 0.05 | 0.04 | 0.02 |
| rash | 0.18 | 0.17 | 0.16 | 0.18 | 0.16 | 0.16 | 0.14 | 0.14 | 0.18 | 0.12 | 0.16 | 0.09 | 0.19 | 0.08 | 0.11 | 0.14 | 0.10 | 0.10 | 0.11 | 0.05 | 1.00 | 0.15 | 0.10 |
| sleep | 0.20 | 0.29 | 0.25 | 0.21 | 0.31 | 0.18 | 0.20 | 0.27 | 0.26 | 0.12 | 0.20 | 0.17 | 0.28 | 0.13 | 0.02 | 0.18 | 0.12 | 0.13 | 0.14 | 0.04 | 0.15 | 1.00 | 0.12 |
| tachycardia | 0.17 | 0.16 | 0.21 | 0.16 | 0.15 | 0.17 | 0.16 | 0.24 | 0.14 | 0.20 | 0.17 | 0.11 | 0.11 | 0.04 | 0.06 | 0.10 | 0.08 | 0.18 | 0.07 | 0.02 | 0.10 | 0.12 | 1.00 |

Table E.4 Legend: Correlation matrix of Pearson's $R^2$ correlation coefficient for all long COVID symptoms included in the model.

#### References

1. Observational Health Data Sciences and Informatics. Standardized Data: The OMOP Common Data Model. 2024. https://www.ohdsi.org/data-standardization/
2. Odysseus Data Services I. ATHENA - OHDSI Vocabularies Repository. 2023.
3. Zhang H, Zang C, Xu Z, et al. Data-driven identification of post-acute SARS-CoV-2 infection subphenotypes. *Nature Medicine*. Jan 2023;29(1):226–235. doi:10.1038/s41591-022-02116-3
4. Al-Aly Z, Xie Y, Bowe B. High-dimensional characterization of post-acute sequelae of COVID-19. *Nature*. 2021;594(7862):259–264. doi:10.1038/s41586-021-03553-9
5. Centers for Disease Control and Prevention. Long COVID or Post-COVID Conditions. https://www.cdc.gov/covid/long-term-effects/?CDC_AAref_Val=https://www.cdc.gov/coronavirus/2019-ncov/long-term-effects/
6. New ICD-10-CM code for Post-COVID Conditions, following the 2019 Novel Coronavirus (COVID-19) (2021).
7. Pfaff ER, Girvin AT, Crosskey M, et al. De-black-boxing health AI: demonstrating reproducible machine learning computable phenotypes using the N3C-RECOVER Long COVID model in the All of Us data repository. *J Am Med Inform Assoc*. Jun 20 2023;30(7):1305–1312. doi:10.1093/jamia/ocad077
8. National Healthcare Safety Network. *COVID-19 Vaccination Modules: Key Terms*. 2025:42–44. https://www.cdc.gov/nhsn/pdfs/hps/covidvax/UpToDateGuidance-508.pdf
9. Centers for Medicare and Medicaid Services. Billing and Coding: Therapy Evaluation, Re-Evaluation and Formal Testing. A53309. DHHS, CMS. Accessed 10/3/2024, 2024. https://www.cms.gov/medicare-coverage-database/view/article.aspx?articleid=53309
10. *MatchIt: Nonparametric Preprocessing for Parametric Causal Inference*. Version 4.7.2. CRAN; 2025.
11. Harrell FE. *Regression Modeling Strategies*. 2nd ed. Springer Series in Statistics. Springer; 2001.
12. Andersen PK, Skovgaard LT. *Regression with Linear Predictors*. Statistics foir Biology and Health. Springer; 2010.
13. Harrell FE. Regression Modeling Strategies. CRAN; 2024.
14. Vatcheva KP, Lee M, McCormick JB, Rahbar MH. Multicollinearity in Regression Analyses Conducted in Epidemiologic Studies. *Epidemiology (Sunnyvale)*. Apr 2016;6(2)doi:10.4172/2161-1165.1000227
15. Kim JH. Multicollinearity and misleading statistical results. *Korean J Anesthesiol*. Dec 2019;72(6):558–569. doi:10.4097/kja.19087
